# Simulation-Based Inference for Whole-Brain Network Modeling of Epilepsy using Deep Neural Density Estimators

**DOI:** 10.1101/2022.06.02.22275860

**Authors:** Meysam Hashemi, Anirudh N. Vattikonda, Jayant Jha, Viktor Sip, Marmaduke M. Woodman, Fabrice Bartolomei, Viktor K. Jirsa

## Abstract

Whole-brain network modeling of epilepsy is a data-driven approach that combines personalized anatomical information with dynamical models of abnormal brain activity to generate spatio-temporal seizure patterns as observed in brain imaging signals. Such a parametric simulator is equipped with a stochastic generative process, which itself provides the basis for inference and prediction of the local and global brain dynamics affected by disorders. However, the calculation of likelihood function at whole-brain scale is often intractable. Thus, likelihood-free inference algorithms are required to efficiently estimate the parameters pertaining to the hypothetical areas in the brain, ideally including the uncertainty. In this detailed study, we present simulation-based inference for the virtual epileptic patient (SBI-VEP) model, which only requires forward simulations, enabling us to amortize posterior inference on parameters from low-dimensional data features representing whole-brain epileptic patterns. We use state-of-the-art deep learning algorithms for conditional density estimation to retrieve the statistical relationships between parameters and observations through a sequence of invertible transformations. This approach enables us to readily predict seizure dynamics from new input data. We show that the SBI-VEP is able to accurately estimate the posterior distribution of parameters linked to the extent of the epileptogenic and propagation zones in the brain from the sparse observations of intracranial EEG signals. The presented Bayesian methodology can deal with non-linear latent dynamics and parameter degeneracy, paving the way for reliable prediction of neurological disorders from neuroimaging modalities, which can be crucial for planning intervention strategies.

## 1. Introduction

Epilepsy is the second most common neurological disease, affecting 50 million worldwide (World Health Organization, 2020). In partial epilepsy, seizures originate in a network of hyperexcitable regions referred to as epileptogenic zone (EZ; Bartolomei et al. (2017)), and then propagate to a secondary connected network, the so-called propagation zone (PZ; Proix et al. (2017)). Drugs are used as a long-term therapeutic treatment, however, 22% to 40% become resistant to medication (Beghi, 2020; Kwan et al., 2010). Resective surgery aims to remove the part of the brain responsible for seizure genesis and is used for such patients (Cardinale et al., 2019). Thus, accurate identification of the EZ/PZ is crucial for planning surgical intervention strategies. While various methods of intracranial EEG recording exist, the method of choice for many epilepsy surgical metrics is stereoelectroencephalography (SEEG; Bancaud et al. (1970)). SEEG consists of invasive implantation of intracerebral electrodes (generally 0.8 mm diameter) including multiple contacts (2 mm long and 1.5 mm apart) targeting different brain areas. The SEEG approach, unlike other intracranial recording methods, allows simultaneous recording from multiple distributed and deeper sites (Cardinale et al., 2016). The outcome of surgery critically depends on the accuracy of the initial hypotheses (e.g., the spatial map of EZ/PZ), and the precision of electrode placement (Bartolomei et al., 2008). Reliable model-based inference on the EZ/PZ from whole-brain epileptic patterns is challenging due to the large dimensionality of the parameter space, non-trivial brain network effects, non-linearity involved in spatio-temporal brain organization, and uncertainties of the model components. Moreover, the dynamics of source states are unobserved, leading to non-identifiability issues for parameter inference using measurements at the sensor-level due to the spatial sparsity of SEEG implantation and source mixing at the sensors. Electromagnetic brain source localization techniques are widely used to reconstruct the underlying cerebral activity from electroencephalographic measurements. However, in the context of SEEG, because of the sparse implantation of electrodes, there is no unique and bijective mapping between the sources and the sensors, rendering the source localization a severely ill-posed inverse problem (Caune et al., 2014; Pizzo et al., 2019).

Bayesian inference is a principled method for updating beliefs with observed data to characterize uncertainty over unknown quantities. This probabilistic methodology provides an estimation of unknown model parameters by incorporating the uncertainty or variation in assumptions, latent variables, measurements, and algorithmic predictions (Bishop, 2006). Bayesian inference from SEEG recordings takes into account the uncertainties in the forward model such as the Virtual Epileptic Patient (VEP; Jirsa et al. (2017)) by allowing constrained variations around model components such as connectome (brain’s anatomical connections), lead-field matrix (mapping the source activities to sensor signals), and epileptogenicity of brain regions (bifurcation parameter). Markov chain Monte Carlo (MCMC; Gelman et al. (1995)) is a non-parametric method that requires explicit evaluation of the likelihood function and is asymptotically unbiased to sample from the posterior distribution (through stochastic transformations). However, evaluation of the target distribution can be prohibitive in high-dimensional spaces, often with the rejection of many proposals that impose the search space exploration to converge very slowly. While Hamiltonian Monte Carlo (HMC; Duane et al. (1987); Neal (2011)) is well suited to sampling from high-dimensional distributions, it may take many evaluations of the log-probability of the target distribution and its gradient for the chain to converge, in particular, when the geometry of the target distribution is unfavorable (Hoffman et al., 2019). To increase the efficiency of HMC sampling for sparse measurements such as SEEG data, more sophisticated reparameterization techniques changing local geometry of the posterior (Betancourt, 2016a, 2014b) are required. Another key disadvantage of directly using the MCMC to fit SEEG data is the lack of amortization. To evaluate evidence for different hypotheses in personalized medicine, the amortization strategies based on artificial neural networks (ANNs) can be immediately applied to arbitrary inference on new recordings without necessitating repeated training (Gershman and Goodman, 2014). After an upfront computational cost at the simulation and training steps to learn all the posteriors, new hypotheses can be evaluated efficiently without computational overhead for further simulations (Cranmer et al., 2020).

Simulation-based inference (SBI) aims to perform flexible and efficient Bayesian inference for complex models when standard methodologies cannot be applied, due to analytic or computational difficulties in calculating the likelihood function (Cranmer et al., 2020). The core of the methodology only requires forward simulations from the computer programming of a parametric stochastic simulator (also referred to as generative model), rather than model-specific analytic calculation or exact evaluation of likelihood function (Beaumont, 2010; Lueckmann et al., 2021; Papamakarios et al., 2019a).SBI is a method for diverse scientific applications where (i) a forward model (simulator) is available, (ii) the likelihood is intractable, and (iii) an accurate approximation with the right amount of uncertainty is important to achieve. In practice, the traditional approximate Bayesian computation (ABC) methods (Beaumont et al., 2002; Sunnaker et al., 2013; Sisson et al., 2018) for posterior estimation suffer from the curse of dimensionality and their performance depends critically on the tolerance level in the accepted/rejected parameter setting (Cranmer et al., 2020; Wrede et al., 2021). An alternative approach is to utilize ANNs to either estimate the posterior directly, bypassing the need for MCMC (Papamakarios and Murray, 2016; Lueckmann et al., 2017; Greenberg et al., 2019), or use synthesized likelihoods or density ratios which require MCMC sampling or training classifiers to extract information from the posterior (Papamakarios et al., 2019b; Hermans et al., 2020; Durkan et al., 2020). Deep neural density estimators form a family of methods that estimate probability densities with the aid of ANNs and have shown impressive performance on challenging problems in scientific fields such as cosmology (Alsing et al., 2018; Zhao et al., 2022), high-energy physics (Brehmer et al., 2018; Brehmer, 2021), and computational neuroscience (Gonçalves et al., 2020; Bittner et al., 2021). These algorithms have reduced the number of simulations needed to perform effective inference by orders of magnitude leveraging ANNs for learning summary statistics. Rather than model simulation during MCMC sampling, ANNs can be trained as parametric function approximators to learn the (log) likelihood function from a set of existing simulations with random parameter values sampled from a prior distribution. After the training step, the likelihood evaluation for new observations only requires a forward pass through the ANNs, with no demand for further simulations at the inference step (i.e., amortized over subsequent evaluations).

Normalizing Flows (NFs) are a family of generative models that convert a simple initial distribution into any complex target distribution, where both sampling and density evaluation can be efficient and exact (Rezende and Mohamed, 2015; Papamakarios et al., 2019a; Kobyzev et al., 2020). NFs leverage ANNs to represent these complex invertible transformations. Here, generative modeling is an unsupervised machine learning method to model a probability distribution given samples drawn from that distribution. In this approach, a simple base probability distribution (e.g., a standard normal) is transformed into a more complex distribution (potentially multi-modal) through a sequence of invertible mapping (implemented by deep neural networks), under the change of variables formula and preserving total probability. It has been shown that NFs systematically outperform other competing approaches for conditional distribution inference and posterior approximation such as amortized variational inference (Rezende and Mohamed, 2015; Papamakarios et al., 2019a). When NFs are conditioned on the observations, they can approximate the posterior distribution very efficiently, and provide all the necessary tools to rapidly sample from the posterior. Note that the output of the classical ANNs is a point estimate of the parameters (or an average value over all data). Rather, NFs transform a simple distribution conditioned on the data to obtain the full probability distribution of the target parameter, even if they are multi-modal (Baso et al., 2021).

The framework we present in this study leverages the power of ANNs to learn the approximate likelihood for efficient and accurate Bayesian inference at the whole-brain level. In particular, the proposed approach used here for parameter inference from SEEG data has several benefits such as (i) no need for source localization or online simulation during inference, (ii) dealing with degeneracy and potential multi-modalities, (iii) training on low-dimensional data features for faster inference of full posterior, (iv) amortized inference at patient-level (i.e., inference on new recording without repeatedly incurring substantial simulation costs). We show that training deep neural density estimators on model simulations is scalable in inferring the whole-brain parameters, and the low-dimensional summary statistics, which retain sufficient information about the parameters of the generative process, can readily provide inference on new data after initial training. We demonstrate the power and flexibility of our approach for the accurate estimation of seizure initiation and propagation from source-level as well as from sensor signals. Using synthetic data, we show that deep neural density estimators can accurately recover posterior parameter distributions at (i) source-level brain activities using only seizure onset or the system fixed point as data feature, (ii) sparse SEEG measurements using only (time-independent) summary statistics. We also investigate the underlying causes of parameter non-identifiability and discuss straight-forward methods for determining when parameters of the model can be estimated uniquely from the sparse observation.

## 2. Materials and methods

### 2.1. Individual patient data

We selected patients initially diagnosed with temporal lobe epilepsy (surgical procedure: resection, surgical outcome: seizure-free, Engel score I). The patients underwent comprehensive presurgical evaluation, including clinical history, neurological examination, neuropsychological testing, structural and diffusion MRI scanning, Stereotactic-EEG (SEEG) recordings along with video monitoring as previously described in Bartolomei et al. (2008); Proix et al. (2017). SEEG electrodes were implanted in the regions suspected to be in the epileptogenic zone. Each electrode had 10-15 contacts (length: 2 mm, diameter: 0.8 mm, contacts separation: 1.5 mm). SEEG signals were recorded with a 128-channel Deltamed system (sampling rate: 512 Hz, hardware band-pass filtering: between 0.16 and 97 Hz). To determine electrode positions, an MRI was performed after electrode implantation (T1 weighted anatomical images, MPRAGE sequence, TR = 1900 ms, TE = 2.19 ms, 1.0 × 1.0 × 1.0 *mm*^3^, 208 slices) using a Siemens Magnetom Verio 3T MR-scanner. To reconstruct patient-specific connectome (DTI-MR sequence, angular gradient set of 64 directions, TR = 10.7 s, TE = 95 ms, 2.0 × 2.0 × 2.0 *mm*^3^, 70 slices, b-weighting of 1000 *s/mm*^2^), diffusion MRI images were also obtained on the same scanner. The study was approved by the Comité de Protection (CPP) Marseille 2, and all patients signed an informed consent form.

### 2.2. Network anatomy

The structural connectome was built with TVB-specific reconstruction pipeline using generally available neuroimaging software (see Fig. 1**A**). The current version of the pipeline evolved from a previously described version (Schirner et al., 2015; Proix et al., 2016, 2017). First, the command *recon-all* from Freesurfer package (Fischl, 2012) in version v6.0.0 was used to reconstruct and parcellate the brain anatomy from T1-weighted images. Then, the T1-weighted images were coregistered with the diffusion weighted images by the linear registration tool *flirt* (Jenkinson et al., 2002) from FSL package in version 6.0 using the correlation ratio cost function with 12 degrees of freedom. The MRtrix package in version 0.3.15 was then used for the tractography. The fibre orientation distributions were estimated from DWI using spherical deconvolution (Tournier et al., 2007) by the *dwi2fod* tool with the response function estimated by the *dwi2response* tool using the *tournier* algorithm (Tournier et al., 2013). Next, we used the *tckgen* tool, employing the probabilistic tractography algorithm iFOD2 (Tournier et al., 2010), to generate 15 millions fiber tracts. Finally, the connectome matrix was built by the *tck2connectome* tool using the Desikan-Killiany parcellation (Desikan et al., 2006) generated by FreeSurfer in the previous step (the patient’s brain is divided into 68 cortical regions and 16 subcortical structures, see Table S1 for label names and indices of sub-divided brain regions). The connectome was normalized so that the maximum value is equal to one (cf. Fig. S1).

**Fig. 1.**
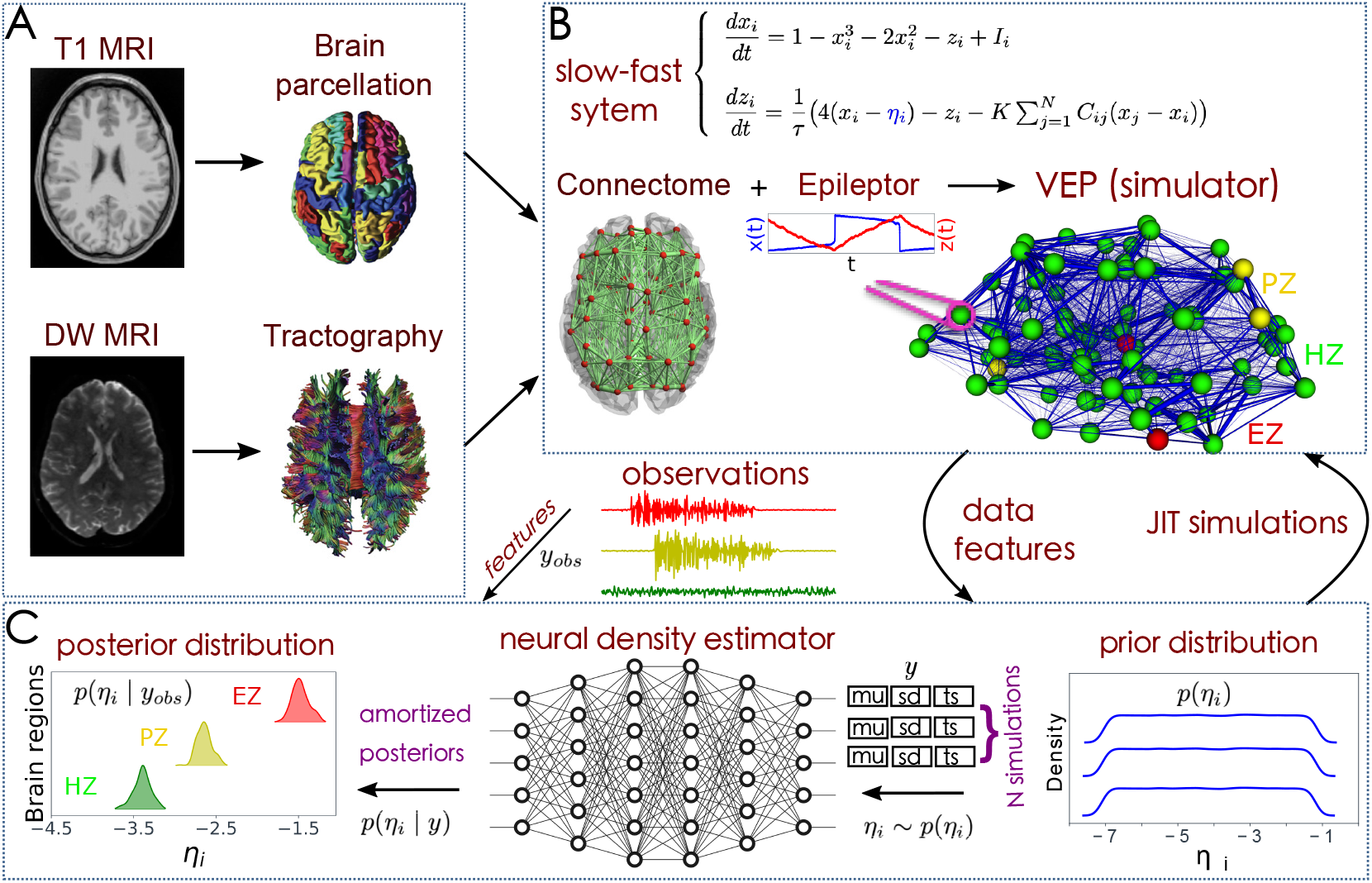
The SBI-VEP workflow to estimate the amortized posterior distribution of the spatial map of epileptogenicity across different brain regions. (**A**) TVB reconstruction pipeline. The T1-weighted MRI images are processed to obtain the brain parcellation. Diffusion-weighted (DW-MRI) images are used for tractography. With the generated fiber tracts and with the regions defined by the brain parcellation, the connectome is built by counting the fibers connecting all regions. (**B**) The Epileptor model as a generic slow-fast dynamical system is merged with patient connectome to build the VEP simulator, which allows the generation of various spatio-temporal patterns. (**C**) SBI with deep neural density estimators. First, the model parameters are drawn randomly from a prior distribution *p*(*η*_*i*_). Then, the VEP simulator takes the parameters as input and generates a simulated dataset as output. A class of Normalizing Flows is then trained on low-dimensional data features of simulations to learn all the (amortized) posteriors *p*(*η*_*i*_ | *y*). Finally, for new observed data *y*_*obs*_, we can readily approximate the true posterior *p*(*η*_*i*_ | *y*_*obs*_).

### 2.3. Stereotactic-EEG (SEEG) data preprocessing

For the selected patients, SEEG electrodes were implanted in critical regions based on the presurgical evaluation.The SEEG data is re-referenced using bipolar montage, which is obtained using the difference of 2 neighboring contacts on one electrode. Bipolar SEEG signal is extracted from 5s before seizure onset up to 5s after seizure offset. The onset and offset times of the epileptic seizure are set by clinical experts. In this study, the log power of high-frequency activity is used as the target for the fitting task. More precisely, the SEEG data are windowed and Fourier transformed to obtain estimates of their spectral density over time. Then, SEEG power above 10 Hz is summed to capture the temporal variation of the fast activity (Jirsa et al., 2017; Proix et al., 2017). The envelope is calculated using a sliding-window approach with a window length of 100 time points. The signal inside the window is squared, averaged and log transformed. Finally, the envelope is smoothed using a lowpass filter with a cut-off in the range of 0.05 Hz. The mean across the first few seconds of the envelope is used to calculate a baseline which is then subtracted from the envelope.

### 2.4. VEP model

To build a whole-brain network model, the brain regions (network nodes) are defined using a parcellation scheme and a set of mathematical equations is placed at each network node to model the regional brain activity (Sanz-Leon et al., 2015; Jirsa et al., 2017). Taking such a data-driven approach to incorporate the subject-specific brain’s anatomical information, the network edges are then represented by structural connectivity (SC) matrix which is obtained from non-invasive diffusion imaging data (dMRI) of the subject (Jirsa et al., 2017; Bansal et al., 2018). Neural mass models are flexible, and physiologically realistic, providing a tractable framework for the analysis of local neural population dynamics by averaging microscopic structure and activity (Jirsa and Haken, 1996; Jirsa et al., 2017; Müller et al., 2020). Neural mass modeling has proven its efficiency in capturing the main features of brain functional behaviors in a single computational framework, by accounting for interactions among brain regions (Jirsa, 2004; David and Friston, 2003; Deco et al., 2009, 2011; Spiegler et al., 2011; Hashemi et al., 2014, 2015; Courtiol et al., 2020).

In the VEP model—a personalized whole-brain network model of epilepsy (Jirsa et al., 2017)—, the dynamics of brain regions are governed by Epileptor model (Jirsa et al., 2014). The Epileptor is a general description of epileptic seizures, which contains the complete taxonomy of system bifurcations to realistically reproduce the dynamics of onset, progression, and offset of seizure-like events (Saggio et al., 2020). The full Epileptor comprises five state variables coupling two oscillatory dynamical systems on three different time-scales: on the fastest time-scale, variables *x*_1_ and *y*_1_ account for fast discharges during the ictal seizure states. On the intermediate time-scale, variables *x*_2_ and *y*_2_ represent the slow spike-and-wave oscillations. On the slowest time-scale, the permittivity state variable *z* is responsible for the transition between the interictal and ictal states. The permittivity variable represents the slow-evolving extracellular processes that occur during epileptiform activity, such as levels of ions, oxygen and energy metabolism, and depending on its values, Epileptor may be driven into or out of a seizure, which accounts for its bi-stable behavior. Additionally, the fast oscillations and spike-and-wave events are coupled via the term *g*(*x*_1_). Following Jirsa et al. (2017), the full VEP brain model equations read as follows:

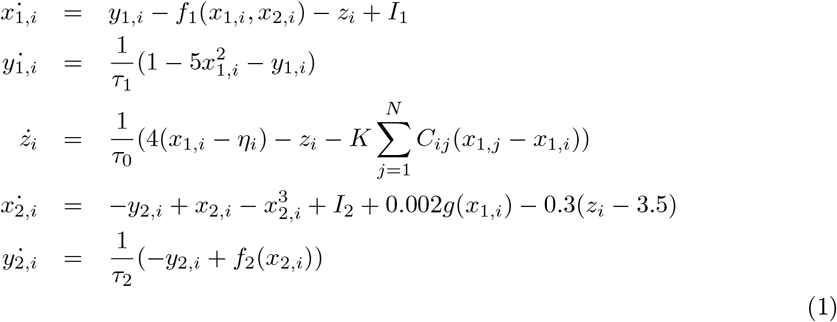

where

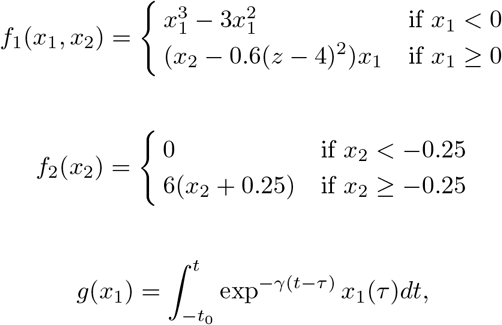

with *τ*_0_ *> τ*_2_ *> τ*_1_. The parameters *I*_1_ and *I*_2_ represent the flow of electric currents that flow inward or outward of neural cells, for the first and second subsystems, respectively. Here *τ*_0_ = 2857, *τ*_1_ = 1, *τ*_2_ = 10, and *γ* = 0.01, *I*_1_ = 3.1, *I*_2_ = 0.45. The degree of epileptogenicity at each brain region is represented through the value of the excitability parameter *η*_*i*_. The network nodes are coupled by a linear diffuse approximation of permittivity coupling through 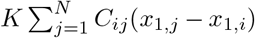, which includes a global scaling factor *K*, and the patient’s connectome *C*_*ij*_.

By applying averaging methods, the effect of the second neuronal ensemble of Epileptor (i.e., the variables *x*_2_ and *y*_2_) is negligible (Proix et al., 2014). Then motivated by Synergetic theory (Haken, 1977; Jirsa and Haken, 1997) and under time-scale separation (*τ*_0_ *≫* 1), the fast variables (*x*_1_ and *y*_1_) rapidly collapse on the slow manifold (McIntosh and Jirsa, 2019), whose dynamics is governed by the slow variable *z*. This adiabatic approximation yields the 2D reduction of VEP model as follows:

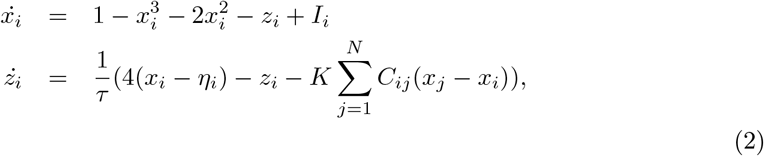

where *x*_*i*_ and *z*_*i*_ indicate the fast and slow variables corresponding to *i*-th brain region, respectively, and the set of unknown *η*_*i*_ is the spatial map of epileptogenicity.

In this study, the full VEP model is used to generate the simulated data, while the Bayesian inversion is based on the 2D reduction of the VEP model to reduce the computational cost associated with the model parameter estimation. The 2D reduction of Epileptor allows for faster inversion while enabling us to predict the envelope of fast discharges during the ictal seizure states (i.e., onset, propagation, and offset of seizure patterns). Additionally, it provides a scheme for inferring slow changes (e.g., extracellular ion concentrations or synaptic efficacy) in biophysical parameters that control fluctuations of fast neuronal states (e.g., seizure activity) (Jafarian et al., 2020, 2021).

### 2.5. Spatial Map of Epileptogenicity

In the VEP, each brain region can trigger seizures depending on its excitability value (node dynamics) and the connectivity to others (network coupling). The parameter *η* controls the tissue excitability, and its spatial distribution is thus the target of parameter fitting. In this study, depending on the excitability parameter, the different brain regions are classified into three main types:

- Epileptogenic Zone (EZ): if *η > η*_*c*_, the brain region can trigger seizures autonomously responsible for the origin and early organization of the epileptic activity. For these regions, the Epileptor exhibits an unstable fixed point, allowing a seizure to happen without a dependency on the network effects.
- Propagation Zone (PZ): if *η*_*c*_ − Δ*η < η < η*_*c*_, the brain region does not trigger seizures autonomously but it may be recruited during the seizure evolution through network effects since its equilibrium state is close to the critical value. For these regions, by a sufficiently large value of external input, a supercritical Andronov-Hopf bifurcation occurs at *η* = *η*_*c*_ corresponding to seizure onset, otherwise, the Epileptor is in its stable equilibrium state.
- Healthy Zone (HZ): if *η < η*_*c*_ − Δ*η*, the brain region stays away from triggering seizures, and all the trajectories in phase-plane are attracted to the single stable fixed point of Epileptor.

Based on the above dynamical properties, the spatial map of epileptogenicity across different brain regions comprises the excitability values of EZ (high value of excitability), PZ (smaller excitability values), and all other regions categorized as HZ (not epileptogenic). Note, however, that an intermediate excitability value does not guarantee that the seizures recruit this area as part of the propagation zone, because the recruitment is also determined by various other factors including structural connectivity, network coupling, and brain state dependence. Using linear stability analysis, isolated nodes displayed a bifurcation at the critical value *η*_*c*_ = −2.05 (Proix et al., 2014; Olmi et al., 2019), and we set Δ*η* = 1.0 (Hashemi et al., 2020).

### 2.6. Simulated Stereotactic-EEG (SEEG) data

Invasive methods such as SEEG implantation are used in clinical situations for patients with drug-resistant epilepsy to determine the focal location of epileptic seizures. The implanted SEEG electrodes record the local field potential generated by the neuronal tissue in its neighborhood. The gain matrix (also known as lead-field matrix or projection matrix) maps the source activity to the measurable sensor signals, as a function of the distance between the sources and sensors. Each sensor collects the source signals in its proximity, weighed by the distance from the sources. A post-SEEG-implantation CT scan is used in this study to find the exact locations of the SEEG electrodes and to calculate the gain matrix. To model the SEEG signals, we assume a linear relation between the source activities and the measurable signals at the sensors:

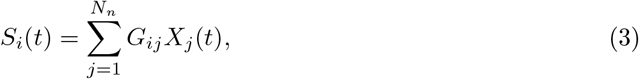

where *S*_*i*_(*t*) is the SEEG signal at sensor *i* ∈ {1, 2, …, *N*_*s*_} with *N*_*s*_ the total number of channels (sensors), *X*_*j*_(*t*) is the source activity in region *j* ∈ {1, 2, …, *N*_*n*_} with *N*_*n*_ the total number of brain regions, and *G*_*ij*_ is the *ij*-th element of the gain matrix mapping the activity at *j*-th region to the *i*-th electrode contact.

Here, the linear combination of source activities (fast variable *x*_1_ in full VEP model given by Eq. (1) or *x* in 2D-VEP model given by Eq. (2)) is governed by the gain matrix whose elements represent the distances of the sensors from the sources. Assuming that the generated signal decays with square of the distance from the source, the gain matrix is approximated by

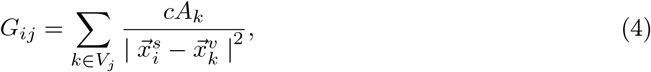

where *V*_*j*_ is the set of all vertices on the triangulate surface of region *j, c* is the scaling coefficient, *A*_*k*_ is the surface associated with vertex *k*, 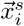 is the position of the sensor *i*, and 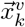 is the position of the vertex *k*. We have not taken into account the dependency of the source-to-sensor decay on the orientation of the neuronal tissue. While the orientation plays an important role for the local field potential generated by the cortical tissue where a clear geometrical arrangement of the neurons exists, it is difficult to quantify this effect for the subcortical structures with their diverse structural arrangements. Thus, due to the lack of information about the orientation in subcortical structures we have chosen to omit the orientation dependency. Note that the sparsity of the gain matrix (cf. Fig. S1**B**) creates difficulties for model inversion in terms of accurate inference of the unknown mixture of activity from different brain regions in the neighborhood of the sensor, computational time, and reliability of the estimated epileptogenicity parameter (structural identifiability).

### 2.7. Generative model

Given a set of observations, the generative model is a probabilistic description of the mechanisms by which observed data are generated through some hidden states (not directly observable) and unknown parameters (not directly measurable). Here, the generative model will therefore have a mathematical formulation guided by the dynamical model that describes the evolution of the model’s state variables, given parameters, over time (Daunizeau et al., 2009, 2014; Hashemi et al., 2020). This specification is necessary to construct the likelihood function (Cooray et al., 2015; Hashemi et al., 2018). The full generative model is then completed by specifying prior beliefs (e.g., dynamical properties and/or clinical knowledge) about the possible values of the unknown parameters (Friston et al., 2014). Notably, the changes in parameter(s) of generative models account for the changes in observations, and the estimation of these parameters enables inference on hidden states that underwrite the changes in observed data, forming the basis for predicting new measurements, causal hypothesis testing, and consensus-based decision making (Friston et al., 2003; Pearl, 2009a; Hashemi et al., 2020). This approach allows us to infer the probability of past and future events, as well as the dynamics of beliefs under changing conditions for generating novel hypotheses where new experiments are prohibitively difficult or impossible to perform (Pearl, 2009b).

In this study, the generative model is formulated on the basis of a system of non-linear stochastic differential equations of the form (so-called state-space representation or evolution equations):

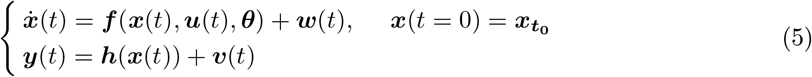

where 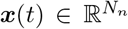 is the *N*_*n*_-dimensional vector of system states evolving over time, 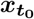 is the initial state vector at time 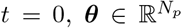 contains all the unknown parameters, ***u***(*t*) stands for the external input, and 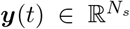 denotes the measured data subject to the measurement error ***v***(*t*). The process (dynamical) noise and the measurement noise denoted by ***w***(*t*) ∼ 𝒩 (0, *σ*^2^) and ***v***(*t*) ∼ 𝒩 (0, *σ*^′2^), respectively, are independent and assumed to follow a Gaussian distribution with mean zero and variance *σ* and *σ*^′^, respectively. Moreover, ***f*** (.) is a vector function that describes the dynamical properties of the system i.e., summarizing the biophysical mechanisms underlying the temporal evolution of system states (here, governed by the VEP model), and ***h***(.) represents a measurement function i.e., the instantaneous mapping from system states to observations (here, the gain matrix).

Considering the 2D reduction of VEP model given by Eq. (2) as the generative model of SEEG recordings, then 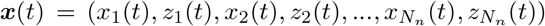, where *N*_*n*_ is equal to the total number of brain regions. By fixing the initial values and the time-scales, 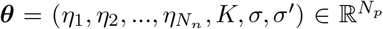, where *N*_*p*_ = *N*_*n*_ + 3. Finally, mean of the observation 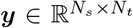 is given by *S*(*t*) = *Gx*(*t*), where 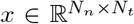 is the fast variable in Epileptor model (cf. Eq. (2)) and 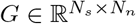is the low-rank gain matrix (cf. Eq. (3)).

### 2.8. Amortized Bayesian inference

In Bayesian framework, the focus is on estimating the entire posterior distribution of the model parameters, i.e., the uncertainty over a range of plausible values for each parameter is naturally quantified, rather than a single point estimate in the Frequentist approach (Hashemi et al., 2018). Bayesian inference is based on the likelihood function of observations given model parameters updated from the prior information (Bishop, 2006). The prior distribution *p*(***θ***) is typically determined before seeing the data through beliefs and previous knowledge about possible values of the parameters, whereas the likelihood *p*(***y*** |***θ***) represents the probability of obtaining the data ***y*** given a certain set of parameter values ***θ*** (the information about the parameters available in the observed data). The likelihood function is typically intractable for high-dimensional models involving non-linear latent variables (such as the VEP model), as it corresponds to an integral over all possible trajectories through the latent space that controls the generative process, i.e., *p*(***y*** |***θ***) = ∫*p*(***y, x*** |***θ***)*d****x***, where *p*(***y, x | θ***) is the joint probability density of data ***y*** and unmeasured latent variables ***x***, given parameters ***θ***. The mapping from each observation ***y*** back to its representation ***x*** in latent space (summarizing the high-dimensional observations) is provided by the generative model (such as the VEP). Once the form of the generative model, i.e., the joint probability distribution of the observation and the parameters *p*(***y, θ***) is defined, we aim to perform inference over the parameters, e.g., using Bayesian framework. By product rule, the generative model can be defined in terms of the likelihood and the prior on model parameters, whose product yields the joint density *p*(***y, θ***) = *p*(***y*** |***θ***)*p*(***θ***). To conduct Bayesian inference, we seek for the posterior distribution *p*(***θ*** |***y***), which is dependent on both prior and likelihood function and can also be used for making predictions about future events (unseen data). Through the Bayes rule, the prior and the likelihood are combined together to obtain the posterior distribution *p*(***θ*** | ***y***):

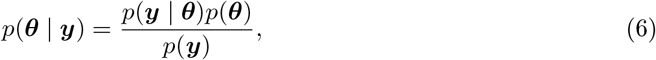

where the denominator *p*(***y***) = ∫*p*(***y, θ***)*d****θ*** = ∫*p*(***y*** | ***θ***)*p*(***θ***)*d****θ*** denotes model evidence (or marginal likelihood, as a quantity of importance for model comparison). This function cannot be explicitly calculated but expressible only as an analytically intractable integral. Thus, the posterior distribution is only known up to a constant of proportionality, and since model evidence is a function of only the data, in the context of inference amounts to simply a normalization term.

Although Markov chain Monte Carlo (MCMC; Gelman et al. (1995); Bishop (2006)) is the most common class of algorithms used in Bayesian analyses for asymptotically exact inference (in the limit of long/infinite runs), there are alternative algorithms to approximate probability distribution. Variational inference (VI; Jordan et al. (1999); Wainwright and Jordan (2008)) is a widely used technique to approximate posterior distributions via simpler approximating distributions. VI turns the Bayesian inference into an optimization problem, which typically results in much faster computation than MCMC methods (Gelman et al., 1995; Kucukelbir et al., 2017). Despite the success and ongoing advances in improving the performance of variational families, there are several constraints on these approximation techniques that limit their power as a default method for statistical inference (Rezende and Mohamed, 2015). For instance, the standard variational method in the Variational Autoencoder uses independent univariate normal distributions to represent the variational family. The popular mean-field approximation assigns an approximating variational distribution to each parameter independently. However, the true posterior in practice is neither independent nor normally distributed, which restricts us from not being able to infer general elliptically symmetric or multi-modal distributions with heavy or light tails (MacKay, 2003; Blei et al., 2017; Yao et al., 2018).

Since for most types of generative whole-brain models, the exact evaluation of likelihood function (integration over the latent variables) is often intractable (either it does not have closed-form expression, or it is computationally prohibitive to obtain), here, we use approximate inference schemes that rely on the use of deep neural networks. Our choice is motivated by the theoretical and engineering advances making the task of training generative models significantly more approachable than in the past. More specifically, we aim to find a parametric density family *q*_*ϕ*_(***θ***) over a shared set of variational parameters *ϕ*, that for a given ***θ***, best approximate the actual posterior *p*(***θ*** | ***y***).

Normalizing Flows (NFs) (Tabak and Turner, 2013; Rezende and Mohamed, 2015; Papamakarios et al., 2019a; Kobyzev et al., 2020) is a family of methods for constructing any complex probability distribution from a simple distribution through a chain (flow) of invertible (bijective), differentiable (smooth), and parametric transformations, often implemented by ANNs. Let ***u*** ∈ ℝ^*d*^ be a random variable and ***f*** : ℝ^*d*^ → ℝ^*d*^ an invertible smooth mapping with inverse ***f*** ^−1^. We can use ***f*** to transform random variable ***u*** with distribution *p*_*u*_(***u***). By applying the change of variables formula from probability theory, the resulting random variable ***u***^***′***^ = ***f*** (***u***) has the following probability distribution:

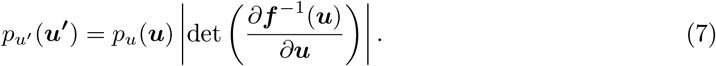

We can construct arbitrarily complex densities (i.e., non-Gaussian) by composing several simple maps and successively applying the transformation as given by Eq. 7. The defining property of flow-based models is that the transformation ***f*** must be invertible and both ***f*** and ***f*** ^−1^ must be differentiable (i.e., diffeomorphisms).

NFs approximate the true posterior distribution *p*(***θ*** | ***y***) from a base probability distribution *p*_*u*_(***u***) by applying diffeomorphism transformation ***θ*** = ***f*** (***u***). If a NF is able to learn the mapping between a simple prior distribution and a complex posterior distribution, the inverse transformation enables us to sample from the posterior by simply extracting values from the prior distribution and applying the learned transformations (Baso et al., 2021). By preserving total probability, and applying the change of variables formula, the posterior *p*(***θ*** | ***y***) is given by

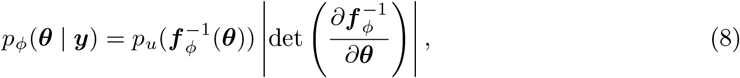

where the first factor represents the probability density for the base distribution *p*_*u*_ evaluated at 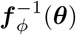, and the second factor is the absolute value of the Jacobian determinant which accounts for the change in the volume due to the transformation. Generally, the main bottleneck in using the change of variables formula is computing the determinant of the Jacobian. An important property of diffeomorphisms is that they are composable, and for such transformations, their composition is itself a diffeomorphism (Papamakarios et al., 2019a). If we compose a chain of K transforms ***f*** = ***f*** _1_ *∘****f*** _2_ *∘· · ·∘* ***f*** _*K*_, their inverse can also be decomposed in the components 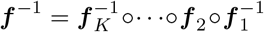 and the Jacobian determinant is the product of the determinant of each component. We can apply a sequence of diffeomorphisms ***f*** _*k*_ with a finite number of simple transformations *k* ∈ 1, 2, …, *K* ∈ ℕ^+^ to obtain a NF:

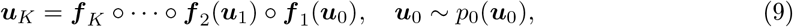

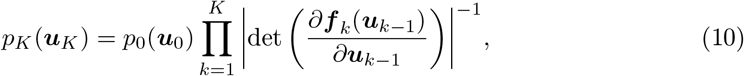

where ***u***_*k*_ = ***f*** _*k*_ (***u***_*k*−1_), with the initial distribution ***u***_0_. In terms of functionality, the transformation must be flexible and expressive enough to model any desired distribution with computational efficiency (i.e., calculating both forward and inverse transformations and associated Jacobian determinants needs to be tractable and efficient). If the transformations are conditioned on observations, the NFs can be trained to return Bayesian posterior probability estimates for any observation. Using NFs, first a sample is drawn from a base distribution, the sample is then transformed with a number of flows, and after applying K flows, the corresponding log-probability of the overall transformation is then approximated by

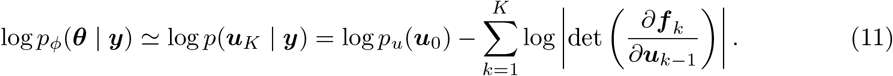

ANNs are often used as inspiration for finding effective transformations. Among many designed architectures for constructing transformation to model high-dimensional complex distributions (NICE; Dinh et al. (2015), Real-NVP; Dinh et al. (2017), PixelRNN; Van Oord et al. (2016), WaveNet; Oord et al. (2016)), Mixture Density Network (MDN; Dockhorn et al. (2020)), and Neural Spline Flows (NSFs; Durkan et al. (2019)), we focused on Masked Autoregressive Flow (MAF; Papamakarios et al. (2017)), which supports invertible non-linear transformations, and enables highly expressive transformations. In MAF, the transformation layer is built as an autoregressive neural network for low-cost computation of the determinant, thus, fast training, and fast to evaluate once trained. This class of deep neural density estimators has achieved state-of-the-art performance as shown to efficiently represent rich structured, and multi-modal posterior distributions (Papamakarios et al., 2019a). Autoregressive flows are universal approximators as they can represent any function arbitrarily well with enough computing (Papamakarios et al., 2019a; Kobyzev et al., 2020). The autoregressive constraint is a way to model sequential data since each output depends only on the data observed in the past (but not on the future ones), whereas the masked conditioners (binary matrices) eliminate the need for the sequential recursion in the ANNs, thus makes MAF fast to evaluate and train on parallel computing architectures (Papamakarios et al., 2017). Due to the autoregressive structure, the Jacobian is triangular by design, hence its absolute determinant can be easily obtained. This property allows us to factorize a target density as a sequence of simpler conditional densities and decompose the joint density into a product of one-dimensional conditional densities according to the probability chain rule. Each conditional probability is modeled by a parametric density, of which the parameters are learned by neural networks. MAF generates each data conditioned on the past dimensions, and density estimation only needs a single forward pass through the flow using architecture like MADE (Germain et al., 2015).

For the purpose of fitting the VEP model to the brain epileptic patterns, we trained NFs by maximizing the log-likelihood of the observed data under our model (Eqs (2) and (3)), equivalently, minimizing the Kullback-Leibler divergence or discrepancy between the true posterior distribution *p*(***θ*** | ***y***) and the variational approximation of Eq. (11) denoted by a family of densities *q*_*ϕ*_, through learning the variational parameters *ϕ*. In practice, this is performed by adjusting the weights *ψ* of a neural network *F* so that *q*_*F* (***y***,*ψ*)_(***θ***) *≃ p*_*ϕ*_(***θ*** | ***y***). Assuming that our dataset has *N* samples 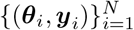 with parameters drawn from the prior *p*(***θ***) and observations generated from the forward model *p*(***y*** | ***θ***), the posterior *p*(***θ*** | ***y***) is obtained by minimizing the loss function

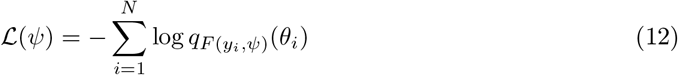

over network parameters *ψ*. After the parameters of the neural networks are optimized, for new observed data ***y***_*obs*_, we can efficiently estimate the target posterior *p*(***θ*** | ***y***_*obs*_) by 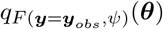. This approach systematically outperforms other competing approaches for posterior approximation by providing a tighter variational lower bound to the marginal log-likelihood (Rezende and Mohamed, 2015). For sufficiently expressive *F* and *q*, we can construct distributions that are more complex than the base distribution and yet have easy sampling and computationally tractable evaluation.

### 2.9. Simulation-based inference (SBI)

Typically, simulators (i.e., a set of dynamical equations such as the VEP) implement a stochastic generative process based on a mechanistic model to produce output through a series of latent states. For many high-dimensional dynamical models that involve expensive computations such as integrals, the calculation of likelihood of the observed data given parameters can become intractable, rendering the likelihood-based inference approaches inapplicable. In this case, the simulator can be used as a black-box whose internal workings are not accessible and are not required to be differentiable, but it can generate synthetic data similar to the observed (empirical) data, allowing to make inferences without access to the likelihood function. Inferring the model parameters from a low-dimensional representation of synthetic data to bypass the evaluation of the likelihood function is often referred to as approximate Bayesian computation (ABC; Beaumont (2010)), likelihood-free inference (LFI; Papamakarios et al. (2019a)), or simulation-based inference (SBI; Cranmer et al. (2020)). In Bayesian terminology, SBI enables us to approximate the posterior distribution of parameters of interest conditioned on observed data with the aid of only forward simulations and avoiding computing potentially intractable log-likelihood and its gradient. Given a prior over parameters, a stochastic simulator, and the observations, SBI returns the posterior distribution that best explains the data.

The classical ABC approaches require the design of a distance metric on summary features, as well as a rejection criterion (*ϵ*), and are exact only in the limit of *ϵ* → 0 (i.e., many rejections). However, in practice, ABC-related methods suffer from the curse of dimensionality, scale poorly to high-dimensional and non-Gaussian data, and are sensitive to the ad-hoc choices (i.e., rejection thresholds, distance functions, and summary statistics), which significantly affect both the computational efficiency and accuracy (Cranmer et al., 2020). Sequential sampling methods using neural network-based density estimators for SBI solve these issues. These algorithms can be divided into three categories referred to as (i) sequential neural posterior estimation (SNPE; Papamakarios and Murray (2016); Lueckmann et al. (2017); Greenberg et al. (2019)), (ii) sequential neural likelihood estimation (SNLE; Papamakarios et al. (2019b); Lueckmann et al. (2019)), (iii) sequential neural ratio estimation (SNRE; Hermans et al. (2020); Durkan et al. (2020)).

SNPE methods refine the estimated posterior in the spirit of adaptive sampling from a simulator, conditioned on a target observation. They directly estimate the posterior density, which can also be used sequentially to train across multiple rounds (the posterior of the previous round is the next proposal prior), in order to reduce the number of calls to the simulator. SNPE family has been shown to perform well across a variety of test problems (Lueckmann et al., 2021). Several SNPE methods have been developed recently based on different choices of the loss function for later rounds, namely SNPE-A (Papamakarios and Murray, 2016), SNPE-B (Lueckmann et al., 2017), and SNPE-C (Greenberg et al., 2019). In this paper, we used SNPE-C (or automatic posterior transform) as it avoids a post-hoc analytical correction and importance weighted loss, which can have high variance during training, leading to inaccurate inference (Lueckmann et al., 2021). By dynamically reparameterizing the proposals and the formulation of the loss function for training, it recovers the true posterior directly (Greenberg et al., 2019). The PyTorch-based SBI package (Tejero-Cantero et al., 2020) implements state-of-the-art algorithms for neural network-based density estimation based on sequential sampling methods (SNPE/SNLE/SNRE). The SBI package works with any simulator as long as it can be wrapped in a Python callable, with a flexible choice of network architectures (Tejero-Cantero et al., 2020; Gonçalves et al., 2020). The simulation step can be easily run on multiple CPU/GPU cores to benefit from effective computational parallelization, and the likelihood function is encapsulated as a feed-forward ANN allowing for parallel evaluation by design, referred to as likelihood approximation networks (LANs; Fengler et al. (2021)).

To perform SBI using SNPE, three inputs are needed to be specified (Gonçalves et al., 2020): (i) a prior distribution describing the possible range of parameters, and we can draw samples from it easily, (ii) a mechanistic model as a simulator that takes parameters as the input and generates simulated data as the output, (iii) a set of observed data (or low-dimensional data feature) as the target of fitting.

Taking prior distribution *p*(***θ***) over the parameters of interest ***θ***, a limited number of simulations (*N*) are carried out to generate a dataset 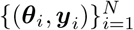, where ***θ***_*i*_ ∼ *p*(***θ***) and ***y***_*i*_ is simulated data given model parameters ***θ***_*i*_. In other words, the simulated data set is a set of *N* independent and identically distributed samples from the generative model *p*(***θ, y***) = *p*(***θ***)*p*(***y*** | ***θ***). Then, we estimate the posterior *q*_*ϕ*_(***θ*** | ***y***) by training the NFs on the generated data set 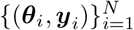. Once the distribution is learned, for new observed data ***y***_*obs*_, we can readily approximate the true posterior *p*(***θ*** | ***y***_*obs*_). Active learning can be used in this approach to adaptively reduce the number of simulations (over multiple simulation rounds by adding their loss terms together (Lueckmann et al., 2019)). In particular, SNPE-C dynamically refines the proposals, network weights, and posterior estimates to learn how model parameters are related to observed summary statistics of the data (Greenberg et al., 2019). Using low-dimensional sufficient statistics, this approach substantially speeds up inference even for models where likelihood can be obtained via numerical integration but with substantial cost. Moreover, no further simulations or training is necessary to estimate the posterior of a new observation when the ANNs training is amortized.

### 2.10. Data features

For many dynamical models, the simulation output is high-dimensional, and the summary statistics are used as a dimension reduction technique for faster training (Sisson et al., 2018; Wood, 2010; Wrede et al., 2021). Importantly, reducing the high-dimensional data to low-dimensional summary statistics makes inference possible, where the likelihood is intractable. In particular, the choice of informative summary statistics is critical for efficient parameter inference as it determines the similarity/discrepancy between simulated proposals and the observed data. Although SNPE can operate inference without summary statistics, the observations at the whole-brain level (such as SEEG) are high-dimensional. In this study, different summary statistics as data features were extracted, comprising seizure onset/offset, power envelope based on signal energy, and statistical moments calculated independently per channel for each patient.

### 2.11. Evaluation of posterior fit

To measure the reliability of the inference using synthetic data, we evaluate the posterior z-scores (denoted by z) against the posterior shrinkage (denoted by s), which are defined as (Betancourt, 2014a):

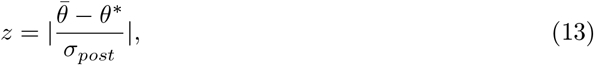

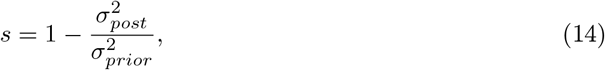

where 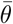 and *θ*^*^ are the estimated-mean and the ground-truth, respectively, whereas 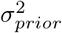, and 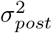 indicate the variance (uncertainty) of the prior and the posterior, respectively. The posterior z-score quantifies how much the posterior distribution encompasses the ground-truth, while the posterior shrinkage quantifies how much the posterior distribution contracts from the initial prior distribution (Betancourt, 2014a). The concentration of estimation towards large shrinkages indicates that all the posteriors in the inversion are well-identified, while the concentration towards small z-scores indicates that the true values are accurately encompassed in the posteriors. Therefore, by plotting the posterior z-scores (vertical axis) against the posterior shrinkage (horizontal axis), the distribution on the bottom right of the plot implies an ideal Bayesian inversion.

### 2.12. Identifiability analysis

In the context of parameter estimation, it is crucial to have knowledge about the identifiability of the parameter estimates, i.e., whether the model parameters can be uniquely determined from the available measurements. Due to the limitation in SEEG implantation in the human brain, only a fraction of the brain regions are close to the electrodes, which could result in non-identifiability issue for the brain regions located far away from the implanted electrodes (i.e., the corresponding elements in the gain matrix are close to zero, see. Fig. S1**B**).

Given a mathematical model describing the observed data measured under specific conditions, different types of non-identifiability can be detected. Structural non-identifiability is only related to the insufficient mapping information from model states to the observables, and it cannot be resolved by increasing the amount or quality of existing measurement conditions. The only remedy for this issue is qualitatively new measurements (e.g., by a change in the placement of SEEG electrodes), which alters the projection mapping (gain matrix). In contrast, practical non-identifiability can arise from a limited amount or quality of recordings under the specific experimental conditions that were used for parameter estimation (e.g., the sampling rate of SEEG recordings or the level of measurement noise). Increasing the amount and/or quality of measured data will ultimately resolve this issue in the estimation. The identifiability analysis is thus required to determine where the model does not yield reliable predictions of system dynamics.

Several approaches such as local sensitivity analysis based on approximated covariance matrix (i.e., the inverse of Hessian or Fisher information matrix) (Rodriguez-Fernandez et al., 2006a,b, 2013), or constructing the confidence regions (Schwaab et al., 2008; Marsili-Libelli et al., 2003; Hashemi et al., 2018) have been proposed to assess the identifiability of the estimated parameters. The uncertainty of estimations in terms of confidence intervals can be assessed by analyzing the actual shape of the likelihood function. For non-linear models and small data samples, the actual shape of the likelihood function typically differs from the standard interval ellipsoid (based on the second order model sensitivities at the optimum), thus, quadratic approximation using the Fisher information matrix can be misleading for identifiability analysis (Raue et al., 2011; Hashemi et al., 2018; Wieland et al., 2021). In particular, for practically non-identifiable parameters, this may result in an incorrect conclusion as structurally non-identifiable (Lill et al., 2019; Wieland et al., 2021). Here, instead of using the local information around the optimum, we use profile likelihood analysis based on the actual shape of the likelihood function, preserving the essential property of boundedness, to assess the identifiability of the estimates. The idea of the approach is to explore the parameter space for each parameter in the direction of the least increase in an objective function (Raue et al., 2011, 2009).

As it is widely known, if we assume that the measurement errors are independent and normally distributed, the maximum likelihood estimation (MLE) and the least-squares estimation (LSE) are equivalent

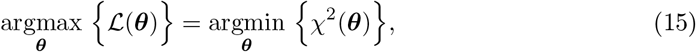

with

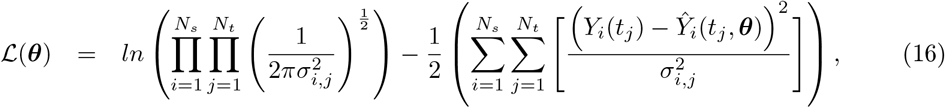

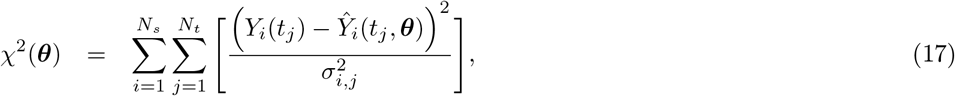

where *ℒ*(***θ***) and *χ*^2^(***θ***) are the log-likelihood and the weighted sum of squared residuals, respectively, *Y*_*i*_(*t*_*j*_) denotes the measured data at *i*-th sensor with *i* ∈ *{*1, 2, …, *N*_*s*_*}* at the time points *t*_*j*_ with *j* ∈ { 1, 2, …, *N*_*t*_}, *Ŷ*_*i*_(*t*_*j*_, ***θ***) represents the corresponding model prediction with ***θ*** as the parameter vector being estimated, and *σ*_*i,j*_ denoting the measurement errors. Here, *N*_*s*_ is the total number of SEEG channels, and *N*_*t*_ is the total number of data points measured per channel. Additionally, if we assume that all variances 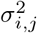 are equal, Eq. (17) simplifies to the well-known chi-squared error criterion (Walter and Pronzato, 1997)

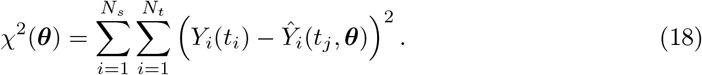

In terms of an objective function such as chi-squared error criterion defined in Eq. (18), the profile likelihood 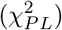 for each parameter individually is defined by

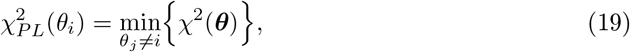

to keep *χ*^2^ as small as possible alongside the fixed *θ*_*i*_, while the minimum for other parameters *θ*_*j* ≠*i*_ is re-obtained for each value of *θ*_*i*_. The identifiability of parameter *θ*_*i*_ is then defined as:

- A structural non-identifiability in the parameter *θ*_*i*_ ∈ *θ*_*sub*_ ⊆ ***θ*** manifests as functional relation 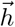 between the parameters *θ*_*sub*_ as:

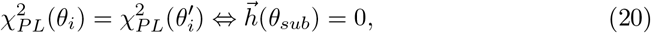

representing a manifold with constant *χ*^2^ in parameter space (*χ*^2^(*θ*_*i*_) = *const*) as *θ*_*i*_ varies without changing the observables, due to the mapping function or the compensation by altering other parameters. The confidence intervals of a structurally non-identifiable parameter are infinite on both sides, hence, its value cannot be uniquely estimated.
- A practical non-identifiability in the parameter *θ*_*i*_ manifests in a likelihood-based confidence interval that is infinite in either the upper or lower bound, although the likelihood has a unique minimum for this parameter. This indicates that the increase in *χ*^2^(*θ*_*i*_) stays below a threshold by increasing and/or decreasing values of *θ*_*i*_.
- A parameter *θ*_*i*_ is identifiable if

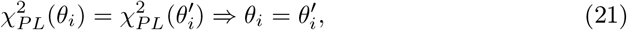

indicating that a unique minimum *χ*^2^(*θ*_*i*_) with respect to *θ*_*i*_ exists with finite confidence intervals in both the upper and lower bounds. The profile likelihood of an identifiable parameter exceeds a threshold for both increasing and decreasing values of *θ*_*i*_.

The threshold Δ_*α*_ = *χ*^2^(*α, df*) to determine the non-identifiability is the *α* quantile of the *χ*^2^-distribution with *df* = 1 degrees of freedom, and *df* = #*θ* being the number of parameters, corresponding to the pointwise and simultaneous confidence intervals, respectively (Raue et al., 2011, 2009).

The identifiability analysis can also be investigated by MCMC sampling methods. From Bayesian perspective, if the posterior distribution follows the prior with no shrinkage, thus, there is no relation between the posterior samples of such parameters, and they are structurally non-identifiable (in a simple example, inference over an extraneous parameter *b* not included in the model *c* = *a*). If there is shrinkage in the posterior but with a statistical relationship (manifold) in the parameter space between the sampled parameters, they are also (structurally or practically) non-identifiable (e.g., inference over parameters *a* and *b* from *c* = *a* + *b* or *c* = *ab*, manifesting as a high correlation in joint posterior samples).

## 3. Results

### 3.1. The SBI-VEP workflow

Fig 1 illustrates the overview of our approach referred to as simulation-based inference for the virtual epileptic patient (SBI-VEP) brain model, which relies only on model-simulations to efficiently estimate the posterior distribution of the spatial map of epileptogenicity, without requiring the exact likelihood evaluation. At the first step to build the SBI-VEP, the non-invasive brain imaging data such as T1-weighted MRI and Diffusion-weighted MRI (DW-MRI) are collected for a specific patient (Fig 1**A**). Using TVB-specific reconstruction pipeline, the brain of the patient is parcellated into different regions (here, using Desikan-Killiany atlas *N*_*n*_ = 84, comprising 68 cortical regions and 16 subcortical structures), which constitute the brain network nodes. The structural connectivity (SC) matrix, whose entries represent the connection strength between the brain regions, is derived from dMRI tractography (see Fig. S1**C**). This step constitutes the structural brain network component, which imposes a constraint upon network dynamics (i.e., the trajectories of the latent state dynamics), as it allows the hidden state dynamics to be inferred from the data. Then, the 2D reduced variant of Epileptor neural mass model (see Eq. (2)) as a generic slow-fast dynamical system is placed at each brain network node and are coupled through SC matrix to reproduce the onset, progression, and offset of epileptic patterns across different brain regions. This combination of the brain’s anatomical information (connectome) with the mathematical modeling of averaged dynamics at the level of local neural populations (e.g., 16 *cm*^2^ of the cortical surface), constitutes the functional brain network model such as VEP model. Then, the VEP simulations can generate various spatio-temporal patterns of whole-brain activity as observed in brain disorders such as epilepsy, with low computational cost (Fig 1**B**). Taking such a whole-brain modeling approach, the spatial map of epileptogenicity across different brain regions (EZ: Epileptogenic Zone, PZ: Propagation Zone, and HZ: Healthy Zone) i.e., parameters *η*_*i*_ ∈ {EZ, PZ, HZ} with *i* ∈ {1, 2, …, *N*_*n*_} is required to be estimated from observations such as SEEG data (see Fig. S1**A**). In the forward modeling of SEEG signals, we assume a linear relation between the non-linear latent dynamics of brain activities at source-level (generated by Epileptor model) and the measured signals at the sensors (see Eq. (3)). Each sensor collects the source signals in its proximity and weighs them by a gain matrix that considers the distance from the sources (see Eq. (4)). Note that the gain (or lead-field) matrix is not of full rank in practice as the number of sources is generally more than the number of sensors due to the sparse placement of SEEG electrodes (see Fig. S1**B**). Finally, we use SBI to estimate the posterior distribution of the VEP model parameters including the brain regional epileptogenicity *η*_*i*_, and global coupling parameter *K* describing the scaling of the brain’s structural connectivity (SC).

The aim of using SBI is to obtain an efficient and accurate approximation to the true posterior of a parameterized stochastic simulator such as the VEP, i.e., dynamical systems from which we can generate samples, but we cannot exactly evaluate the likelihood function. In particular, Normalizing Flows (NFs) embedded in SBI approach such as Sequential Neural Posterior Estimation (SNPE) enable us to approximate the full posterior distribution of parameters conditioned on (low-dimensional summary statistics of) observed data, with the aid of only forward simulations, while also potentially capturing degeneracy or multi-modalities. To perform the SBI, three inputs are needed to be provided: (i) a prior distribution describing the possible range of parameters from which we can easily draw samples, (ii) a simulator in computer code such as the VEP that takes parameters as input and generates data from drawn parameters as output, (iii) a set of low-dimensional data features (sufficient informative statistical summary) for training a neural density estimator. Collecting a simulated dataset by repeatedly drawing samples from the prior distribution specified over parameters and performing model simulations with the randomly chosen parameters, the SNPE trains an ANN such as Masked Autoregressive Flow (MAF) to learn an invertible transformation between data features of simulated dataset and parameters of a parameterized approximation of posterior distribution. After training, SNPE is able to rapidly approximate the full posterior of parameters for new observations or empirical data. By training the deep neural density estimators on a large number of simulations given low-dimensional sufficient statistics, this approach allows for efficient and accurate inference of full posterior, with no demand for further simulations at the inference step due to amortization (i.e., a single pass through the ANNs). A major motivation for this approach is the amortization of parameter inference after the training stage, affording clinicians the ability to evaluate initial hypotheses with negligible computational cost for the inference (in the order of a few seconds).

### 3.2. The SBI-VEP against source-level epileptic patterns

Synthetic data can provide us the ground-truth of EZ/PZ/HZ, which can be used to validate the inference results. As a first example to show the capabilities of the SBI-VEP in the estimation of the spatial map of epileptogenicity, we first fit the epileptic patterns at the source-level time-series of neuronal activity to avoid any non-identifiability issue in the parameter estimation. To this end, we generate a set of observations from the full VEP model (see Eq. (1)) by considering the SC of a patient with partial epilepsy and the gain matrix *G* = *I* as an identity matrix. To generate synthetic observation, we used the Euler-Maruyama integration scheme with time step *dt* = 0.1, global coupling parameter *K* = 1 accounting for interactions among brain regions, and adding an independent zero-mean Gaussian noise with standard deviation of *σ* = 0.1. The seizure initiation and propagation are obtained using the spatial parameters with two regions in EZ (r40: left-hippocampus, r41: left-amygdala) with *η*_*EZ*_ = −1.6, and three regions in PZ (r05: ctx-lh-entorhinal, r06: ctx-lh-fusiform, r32: ctx-lh-temporalpole) with *η*_*P Z*_ = −2.4, while all the other brain regions are set as HZ with *η*_*HZ*_ = −3.65 (i.e., not epileptogenic). In Fig 2**A**, the activity of brain regions in EZ and PZ is shown in red and yellow, respectively. Here, we extract the envelope of signal power (high-pass filtering followed by envelope smoothing) from simulated epileptic patterns, which serves as the data feature for inference. To do this, we used a bandpass Butterworth filter of order 3, with cut-off frequencies between 10 Hz and 100 Hz to remove the baseline shift at seizure onset of observations. Following the Hilbert transformation, a Savitzky-Golay filter with polyorder of 3 was also applied on the analytic signal for the purpose of smoothing in the calculation of the seizure onset. Here we carried out 10^4^ model simulations from the reduced VEP model (see Eq. (2)) to prepare the training data set, while only the statistical moments (*m* = 4) of time-series and the seizure onsets were used as the data features. The parameters are drawn from a uniform prior in the ranges: *η*_*i*_ ∈ *𝒰* (− 5, −1) and *K* ∈ *𝒰* (0, 2), and SNPE was run using a single round. Finally, we used Masked Autoregressive Flow (MAF) to learn the invertible transformation between the feature datasets generated by samples drawn from the prior distribution, and the parameters of approximated posterior distribution. As shown in Fig 2**B**, the predicted time-series are in very good agreement with the observations in terms of seizure initiation, propagation and termination. Note that the observation illustrates the band-pass filtered activity of fast variable in full VEP brain model (i.e., *x*_1,*i*_ in Eq. (1)), whereas the training data set is generated using the simulations from reduced VEP model (cf. Eq. (2)). The estimated posterior distribution of the spatial map of epileptogenicity across different brain regions is shown in Fig 2**C**. From this figure, it can be seen that the true value of *η*_*i*_, with *i* ∈ {1, 2, …, 84} (shown filled black circles) is well under the support of the estimated posterior density across different brain regions. To verify the accuracy and reliability of this result, the posterior z-scores against the posterior shrinkages is investigated on the estimation (cf. Eqs. (13) and (14)). As shown in Fig 2**D**, the distribution of posterior z-scores and posterior shrinkages for all the estimated parameters substantiates reliability of the Bayesian inference using SNPE. The concentration towards large shrinkages indicates that all the posteriors in the inversion are well-identified, while the concentration towards small z-scores indicates that the true values are accurately encompassed in the posteriors. Thus, the distribution on the bottom right of the plot implies an ideal Bayesian inversion. Notably, the estimated posteriors using SNPE are in a very close agreement with those estimated using Hamiltonian Monte Carlo (HMC) algorithm by fitting the envelope of time-series (see Fig. S2). Using SNPE, the only heavy computational task is carrying out the simulations to generate training data. However, this approach is efficient by offering the ability to run model simulations in parallel processors (each VEP run to simulate 100 sec of whole-brain epileptic patterns takes only 0.003 sec using JIT compiler). For SBI, the model simulations can be easily run independently to create the training dataset (the more simulations, the more informative posterior), whereas HMC is amenable to embarrassingly parallel execution of only independent chains as each HMC chain itself is a simulation of Hamiltonian dynamics. Using Masked Autoregressive Flow (MAF) as the deep neural density estimator in SNPE, the posterior estimation took around 1000 sec (for simulation and training as generating 200 samples from posterior took less than 1 sec), whereas generating 200 samples using HMC (with 200 warm-up iterations, the expected acceptance probability of 0.95, and efficient non-centered form of parameterization implemented in Stan (Stan Development Team, 2018)) took around 25000 sec. This indicates that for the VEP model, SNPE is 25 orders of magnitude faster than HMC. Importantly, due to amortized strategy in SBI, we can quickly estimate different EZ/PZ hypotheses without having to carry out new simulations for the inference (by excluding the computational time for training, SNPE is 25000 times faster than HMC). See Fig. S3 for two other analyses with different spatial maps of epileptogenicity across different brain areas. These results indicate the efficiency and accuracy of SNPE that one can efficiently learn all the posteriors of the parameters at whole-brain level from a limited number of model simulations. Moreover, according to our results at sources of epileptic activity, the statistical moments and seizure onset retained adequate information about the parameters of VEP generative process to estimate the spatial map of epileptogenicity. In the following, we investigate the mechanism by which the seizure onset occurs through the changes in the steady-state equilibrium of the system, allowing accurate inference on spatial heterogeneity of model parameters from such low-dimensional but informative data features.

**Fig. 2.**
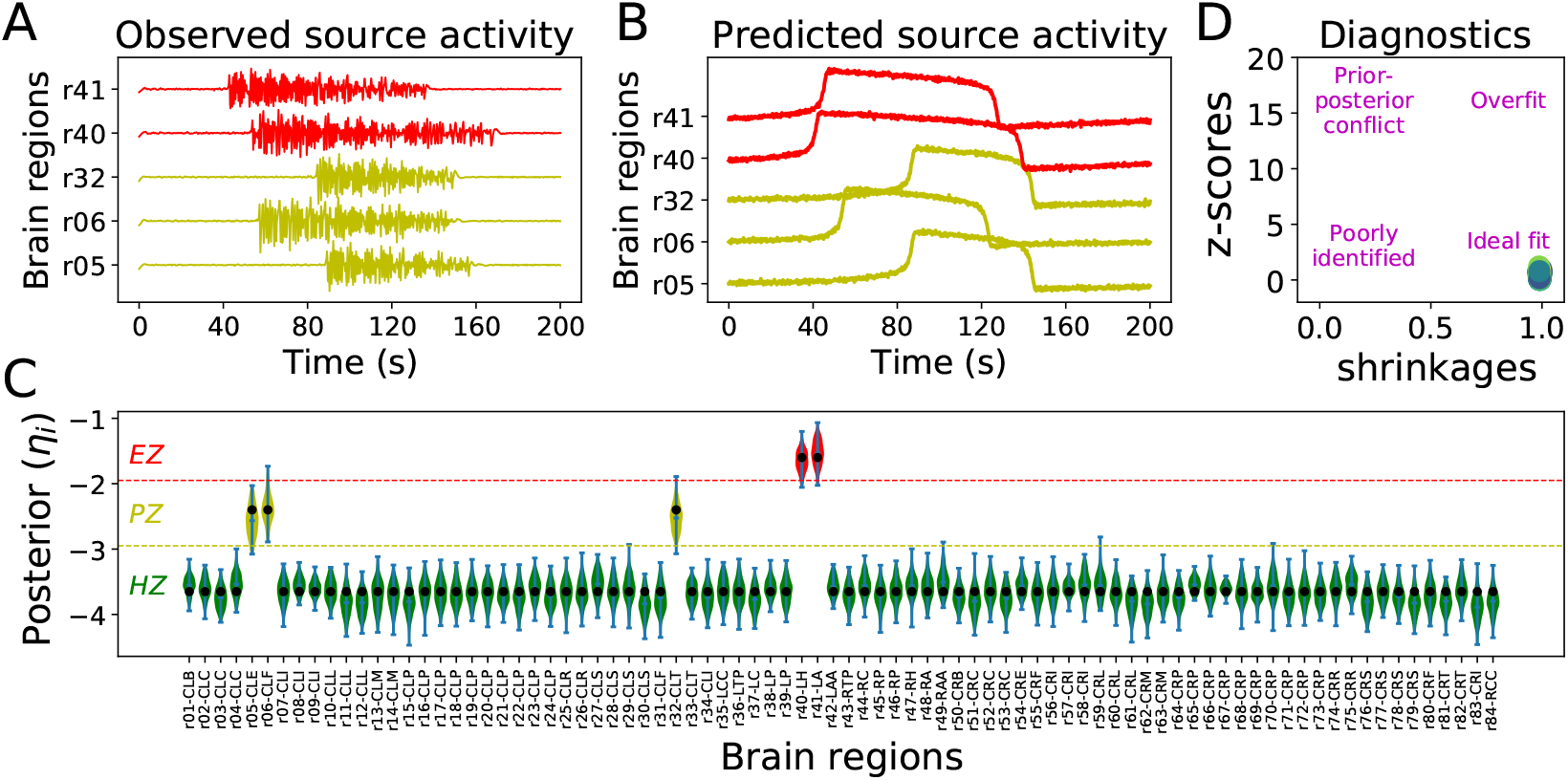
Bayesian inference on spatial map of epileptogenicity across different brain regions using SNPE at source-level brain activity. (**A**) The observed data generated by the full VEP model with two regions as EZ (red), and three regions as PZ (yellow). (**B**) The predicted seizure envelope by training SNPE on a dataset containing 10^4^ simulations of the reduced VEP model, while only statistical moments and seizure onsets were used as the data feature. (**C**) The estimated posterior distribution of *η*_*i*_ for all brain regions. The filled black circles show the ground-truth used to generate the observed data. (**D**) The distribution of posterior z-scores versus posterior shrinkages towards bottom right of the plot implies an ideal Bayesian inversion.

### 3.3. The SBI-VEP against phase-plane dynamics

To illustrate the capabilities of SNPE in capturing the true mechanism underlying seizure initiation and propagation from a dynamical systems theory rather than just data fitting, the phase-plane topology across different brain regions defined as EZ/PZ/HZ is presented in Fig 3. In Fig 3**A**, the observed and predicted brain activity using SNPE at Healthy Zone (HZ) are shown, where all the trajectories in phase-plane settle into the single stable fixed point of the system (not epileptogenic). Fig 3**B** and **C** illustrate the observed and predicted activity at Propagation Zone (PZ), as candidate brain regions responsible for the seizure propagation, in the absence and with sufficiently strong network coupling, respectively. The seizures may be recruited to these regions due to the network effects since their steady-state equilibrium is close to the bifurcation value. This indicates that seizure propagation depends on the interplay between multiple factors including the brain region’s excitability (node dynamics), the individual SC (network structure), the network coupling (parameter *K*), and brain state dependency (such as noise-level). For instance, in the absence of (or with weak) network coupling (e.g., *K* = 0 shown in Fig 3**B**), the seizures cannot spread across the brain, whereas by sufficiently strong coupling (e.g., *K* = 1 shown in Fig 3**C**), the seizure recruitment can be observed in regions defined as PZ. Lastly, Fig 3**D** shows the observed and predicted brain activity using SNPE at Epileptogenic Zone (EZ), where the Epileptor model exhibits an unstable fixed point that allows a seizure to occur as the brain regions’ intrinsic property. This implies that the seizure initiation depends only on the excitability parameter of the brain regions (neither the time-scale of the system, nor the network coupling parameter). From this figure, we observed that SNPE closely follows the true trajectories in the phase-plane that controls the generative process. Notably, we can obtain similar results by training SNPE on the fixed point of the system as the data feature to estimate the spatial map of epileptogenicity in 2D-VEP model (see Fig. S4). Moreover, we analyzed the joint posterior density distributions between all the model parameters (excitability *η*, the system time-scale *τ*, and initial condition of fast and slow variables *x*_*init*_ and *z*_*init*_, respectively, cf. Eq. (2)) for different brain node types as EZ/PZ/HZ (see Fig. S5). SNPE precisely demonstrates a non-identifiability for the system time-scale in HZ, and the initial conditions in EZ and PZ, as expected from dynamical system properties. These results indicate that SNPE is able to accurately identify the stability of an equilibrium in Andronov-Hopf bifurcation from only seizure onset to explain the changing nature of seizure initiation and propagation in a non-linear dynamic system of epilepsy spread.

**Fig. 3.**
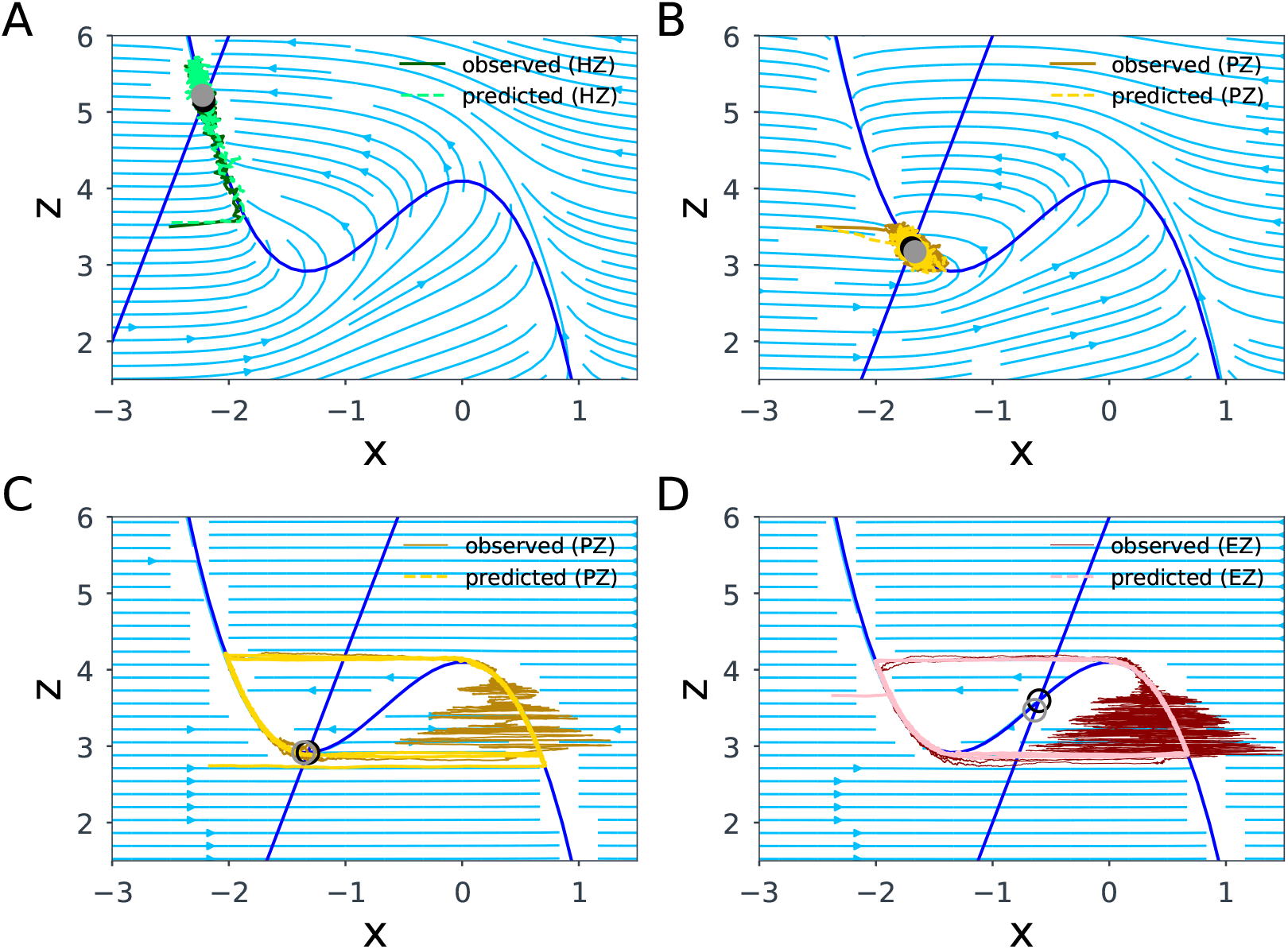
The observed and predicted (non-linear) VEP dynamics using SNPE trained on seizure onsets illustrated in the phase-plane for different brain regions. (**A**) Healthy Zone (HZ) exhibits a single stable fixed point implying that this region is not epileptogenic. (**B**) and (**C**) demonstrating the system dynamics in Propagation Zone (PZ) with the global coupling parameter *K* = 0, and *K* = 1, respectively, as the candidate brain regions for seizure recruitment. Since the equilibrium in PZ is close to the bifurcation value, the seizure spread to these regions is a complex network effect. (**D**) Epileptogenic Zone (EZ) exhibits an unstable fixed point implying that the seizure initiation is a region’s intrinsic property. The intersection of x- and z-nullclines (colored in blue) determines the fixed point of the system. Full circle and empty circle indicate the stable and unstable fixed points, respectively.

### 3.4. The SBI-VEP against SEEG data

Here, we demonstrate the capabilities of the SBI-VEP in the estimation of the spatial map of epileptogenicity from SEEG data, while the seizure dynamics at the source-level of neuronal activity are unknown (i.e., the latent space dynamics). Due to the sparse implantation of electrodes, there is no unique and bijective projection from sensors to sources, consequently, the source localization is a severely ill-posed problem. The sparsity of the gain (lead-field) matrix mapping the sources to the electrode contacts highly impacts the precision of the resulting source reconstruction. Using the generalization (Moore-Penrose) inverse of gain matrix *G*, the pseudo-inverse of matrix *Q* = *G*^⊤^*G* is very sensitive to the tolerance level, i.e., the cutoff for small singular values (see Fig. S6**A**). To avoid this issue, we train SNPE directly on simulated SEEG data at sensor-level to estimate the posterior of the brain regional parameters *η*_*i*_ assuming the lack of knowledge about seizure dynamics (onsets and offsets) at the source space. The aim of using SNPE is to amortized inference on spatial heterogeneity of model parameters from low-dimensional summary statistics while efficiently capturing multi-modalities and dealing with the non-identifiability, thereby avoiding the need for source localization during inference.

Here, the observation is generated from the reduced VEP model (see Eq. (2)) by considering the SC and a low-rank gain matrix *G* of a patient with left partial epilepsy. The ground-truth parameters that generated the observation include the global coupling of *K* = 1, and EZ and PZ networks in the left hemisphere to cover two and three brain regions, respectively, while other brain regions are HZ, according to the clinical hypothesis. A zero-mean Gaussian noise with *σ* = 0.1 was also added to the observation, as shown in Fig 4**A**. To create the training data set, we carried out 10^5^ simulations by sampling from the uniform prior *η*_*i*_ ∈ *𝒰* (−7, −1) and *K* ∈ *𝒰* (0, 2). We trained SNPE with MAF in a single-round, while only the statistical moments (*m* = 4) and the SEEG envelope power (the area under the curve) were used as the data features. As shown in Fig 4**B**, the predicted SEEG signals are in very close agreement with the observation. Fig 4**C** illustrates the estimated posterior of brain regional parameters from low-dimensional summary statistics. We observe that for the regions close to electrodes (including EZ and PZ), the posterior is tightly centered on the ground truth parameters, whereas for many regions that are distant from the electrodes, there is no shrinkage in the estimated posterior from the defined prior (see also Fig. S6**B**). This is due to the zero elements in gain matrix that conceal any impact from the activity of these regions on the observed SEEG data. The poorly-identified excitability for the distant regions from electrodes due to their non-identifiability results in the distribution of posterior z-scores versus posterior shrinkages to have fat tails towards lower shrinkage, as shown in Fig 4**D**. The posterior distribution for the global coupling parameter *K* is centered around the ground truth used to generate the observation, with a proper amount of uncertainty (see Fig. S6**C**). The posterior predictive check (i.e., generating data from the model using the parameters drawn from the estimated posterior, then comparing with observed data) validates the reliability of SNPE, as it correctly fits the target SEEG envelope power (see Fig. S6**D**). Importantly, due to the amortized strategy, we can carry out fast Bayesian inference for arbitrary observations without the need for further simulations. See Fig. S7 for inference results on two other hypotheses about the spatial distributions of excitability parameters. These results indicate the efficiency and accuracy of SNPE to infer brain spatial parameters from SEEG data without access to the knowledge about the brain activities hidden at source-level data. For the sake of comparison with HMC, our non-centered form of model parameterization over state transition probabilities failed to converge to the desired distribution (using Stan with 2000 warm-up and 2000 sampling iterations and the expected acceptance probability of 0.95). HMC can deal with moderate degeneracy, but with extensive use of computational resources. Due to the sparsity of gain matrix and the consequent non-identifiability issue, HMC exhibited undesirable random walk behavior, and demanded a very small value of step-size (e.g., 10^−18^) in leapfrog integration during Hamiltonian dynamic simulation. To efficiently fit SEEG data with HMC, a more sophisticated reparameterization over model configuration space is required for convergence to the typical set. Identifiability analysis on brain region estimation continues this discussion in the next subsection.

**Fig. 4.**
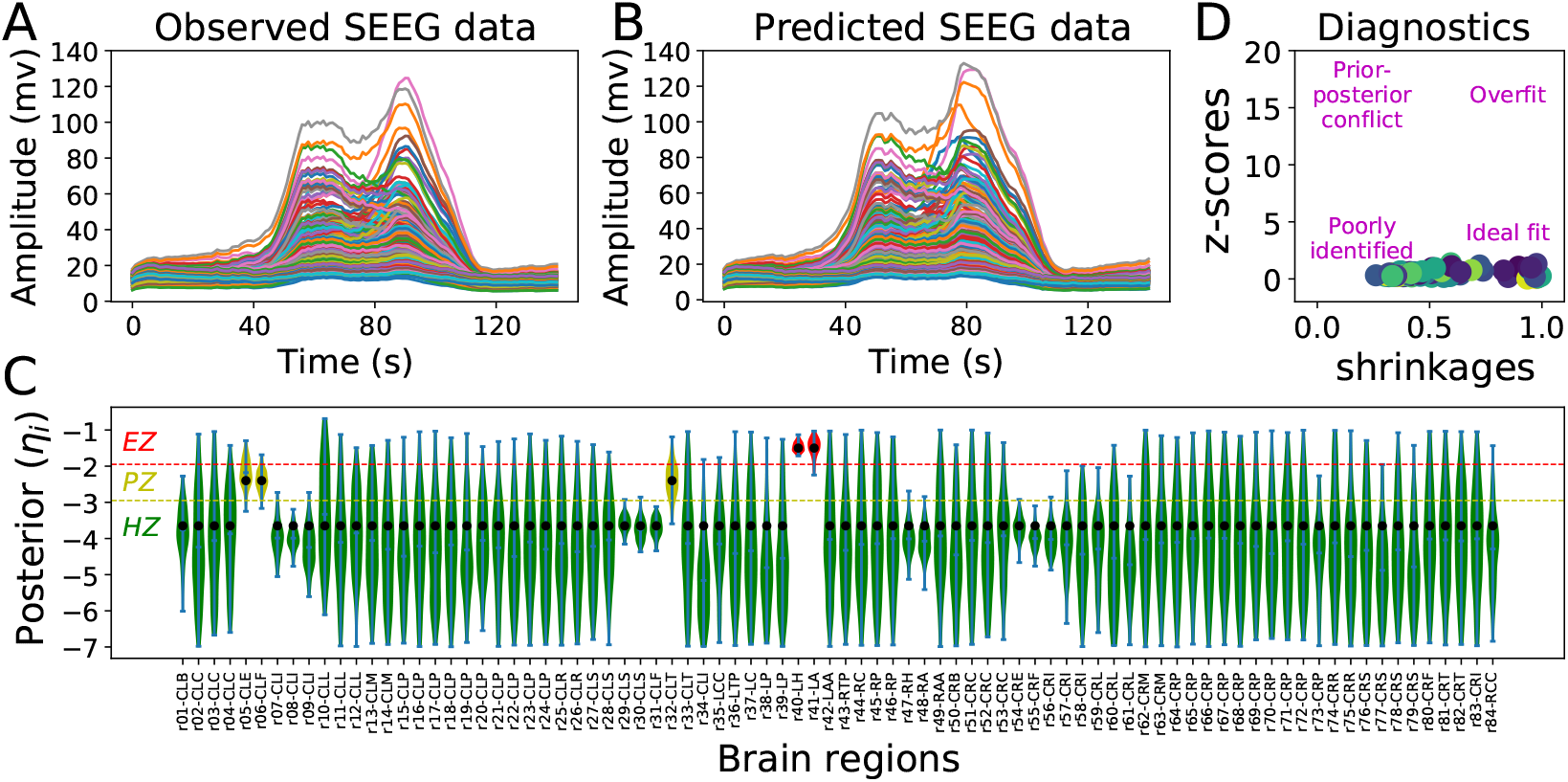
Bayesian inference on spatial map of epileptogenicity across different brain regions using SNPE from SEEG signals. (**A**) The observed SEEG data with two regions in EZ, and three regions in PZ. (**B**) The predicted SEEG data by training SNPE on a dataset generated by the reduced VEP model, while only time-independent summary statistics were used as the data feature. (**C**) The estimated posterior distribution of *η*_*i*_ for all brain regions (green: HZ, yellow: PZ, red: EZ). The filled black circles show the ground-truth used to generate the observation. (**D**) Although the regions close to electrodes are well-identified, the zero elements in the gain matrix (due to sparse implantations) impede an ideal Bayesian inversion at whole-brain level. The poorly-identified excitability for the distant regions from electrodes results in the distribution of posterior z-scores versus posterior shrinkages to have fat tails towards lower shrinkage.

### 3.5. Identifiability analysis in the SBI-VEP

In the context of parameter estimation for large-scale brain networks, it is essential to investigate the identifiability of the regional parameters, i.e., whether the excitability parameter across different brain regions can be uniquely determined from the available measurements. From a Frequentist point of view, insufficient information in the observed data can be interpreted as non-identifiability. The profile-likelihood approach offers a robust way to detect both structural and practical types of non-identifiability from the actual shape of likelihood function. Here, we first demonstrate the parameter identifiability analysis using likelihood-based approach, which relies on confidence intervals of the maximum-likelihood estimates (MLEs, see Eq. (16)). The profile likelihood (see Eq. (19)) is calculated by iteratively shifting one parameter from its optimal value by a small change, followed by fitting again the other parameters in the model. Using this approach, parameter *η*_*i*_ is identifiable, if a unique minimum of *χ*^2^(*η*_*i*_) exists, and the profile likelihood reaches both an upper and lower threshold, thus leading to finite confidence intervals, as shown in Fig. 5**A**. When the quantile *α* = 0.95, the identifiability threshold is Δ_*α*_ = 3.84. In two-dimensional parameter space (e.g., two regions close to electrodes with different distances), this is demonstrated as a valley with finite confidence intervals. If a unique minimum exists but with a finite confidence interval in only upper or lower bound, the parameter is practically non-identifiable. In two-dimensional parameter space (e.g., two regions close to electrodes with same distance), this can be visualized as a relatively flat valley, which is infinitely extended in one direction, as shown in Fig. 5**B**. The practical non-identifiability arises due to the limited amount and/or quality of observations. The remedy for this issue is to add sufficient data to the existing observables such that it provides sufficient constraining power in latent space dynamics, yielding a unique estimation with finite confidence intervals. The likelihood-based confidence intervals of a structurally non-identifiable parameter are infinite in both the upper and lower bounds, hence they cannot be uniquely estimated from observation. For a pair of the structurally non-identifiable parameters (e.g., two distant regions from electrodes), this results in a flat valley, infinitely extended along with the corresponding functional relation, as illustrated in Fig. 5**C**. No single combination of parameter values on this surface can provide a single best fit to the data. Rather, a vast contour through the parameter space fits the data equivalently. Since structural non-identifiability is independent of the accuracy of experimental data, this issue cannot be resolved by increasing the amount and/or quality of existing measurements, even as we collect an infinite number of repeated observations. The only remedy for this issue is designing a new setup for the measurements (such as new SEEG implantation, which alters the mapping function through the gain matrix), or a reparameterization over model configuration space.

**Fig. 5.**
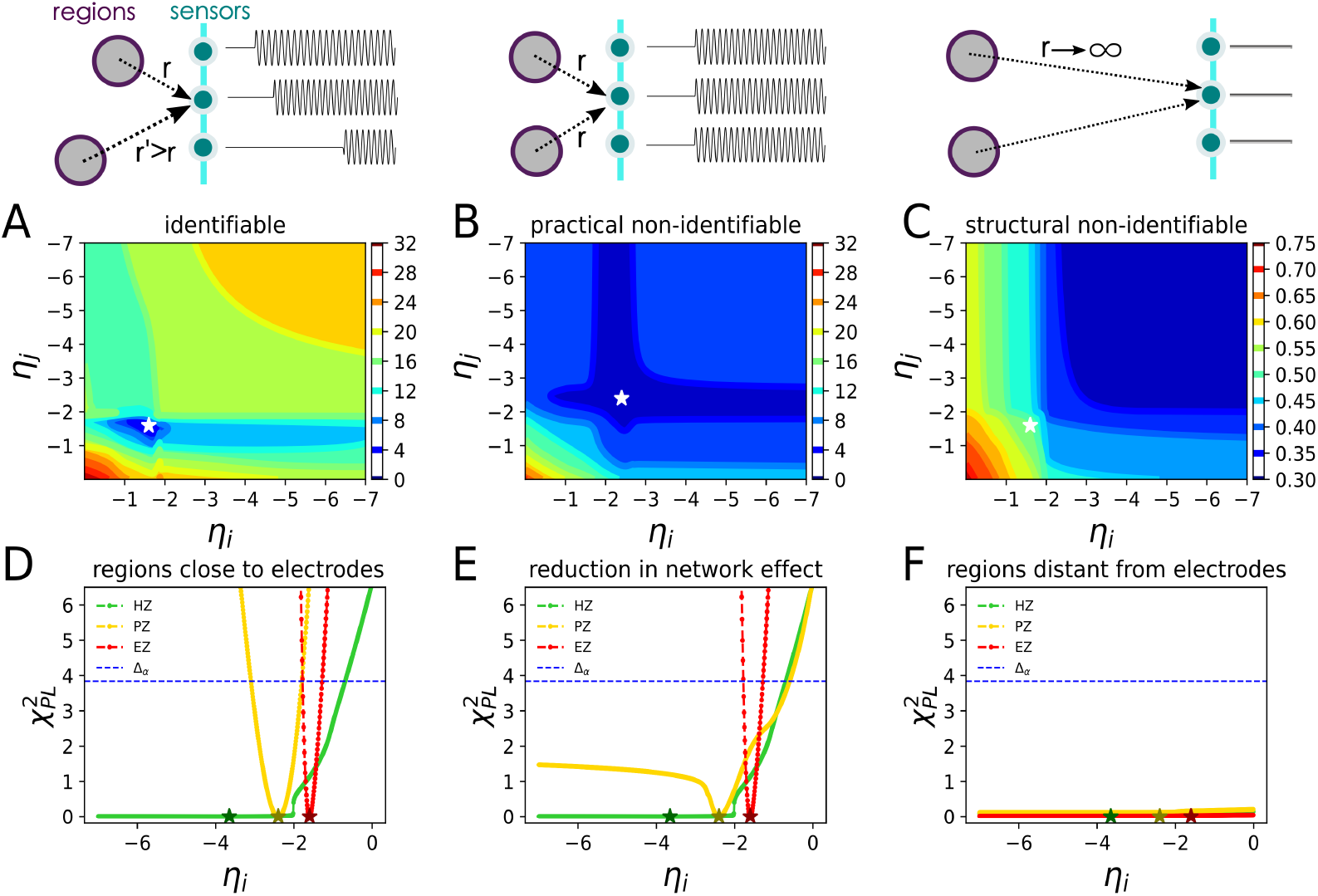
Frequentist identifiability analysis in the SBI-VEP. (**A**), (**B**), and (**C**) show the 2D contour plots of profile likelihood for identifiable (two regions close to electrodes with different distances), practically non-identifiable (two regions close to electrodes with same distance) and structurally non-identifiable (two distant regions from electrodes) parameters, respectively. Contour lines colored from blue to red correspond to low to high values of profile likelihood, respectively. The white asterisk indicates the ground-truth. (**D**), (**E**), and (**F**) show the profile likelihood for the brain regions close to electrodes, by a reduction in patient’s SC, and for the brain regions located far away from electrodes, respectively. When *α* = 0.95, the identifiability threshold is Δ_*α*_ = 3.84 (horizontal dashed blue line). According to profile likelihood, identifiable: finite confidence intervals, practically non-identifiable: finite confidence interval in only upper bound, structurally non-identifiable: infinite confidence intervals in both directions.

Based on the profile likelihood for the near-electrode regions indicates that the EZ and PZ are identifiable, but the HZ is practically non-identifiable, since its steady-state equilibrium is far from the bifurcation value. See Fig. 5**D**, showing that the profile likelihood of EZ and PZ is constrained in both upper and lower bounds (in red and yellow parabolas, respectively), whereas for HZ (in green), only a finite upper confidence bound is derived at the bifurcation value *η*_*c*_ = −2.05. Notably, by a 50% reduction in the global coupling parameter *K*, the EZ and HZ remained unchanged in terms of identifiability (Fig. 5**E**, in red and green, respectively), implying that the activity in these regions is a local property. However, the PZ now recasts as a practically non-identifiable (Fig. 5**E**, in yellow), indicating that the seizure recruitment to PZ is a network effect. For the distant regions from the electrodes, all EZ/PZ/HZ manifest as structurally non-identifiable by the virtue of a flat profile likelihood in any direction (Fig. 5**F**), indicating that no unique solution exists for MLEs. In the shown panels, the asterisks denote the ground-true values, and the dashed blue lines denote the threshold Δ_*α*_ = 3.84 that yields a likelihood-based confidence region.

In terms of reliability in SNPE estimation for identifiable and non-identifiable regions, the uncertainty in estimated posterior distributions is illustrated in Fig. 6**A** and **D**, respectively, while the node dynamic is varied by the change in the excitability parameter. For an identifiable region, the estimation by SNPE demonstrates an accurate and robust inference by recovering the ground truth with a proper amount of uncertainty, even if the number of electrodes is more than the number of regions (Fig. 6**A**). However, for a non-identifiable region (distant from electrodes), the estimation by SNPE results in a high amount of uncertainty, as the excitability parameter cannot be determined uniquely for such regions (Fig. 6**D**).

**Fig. 6.**
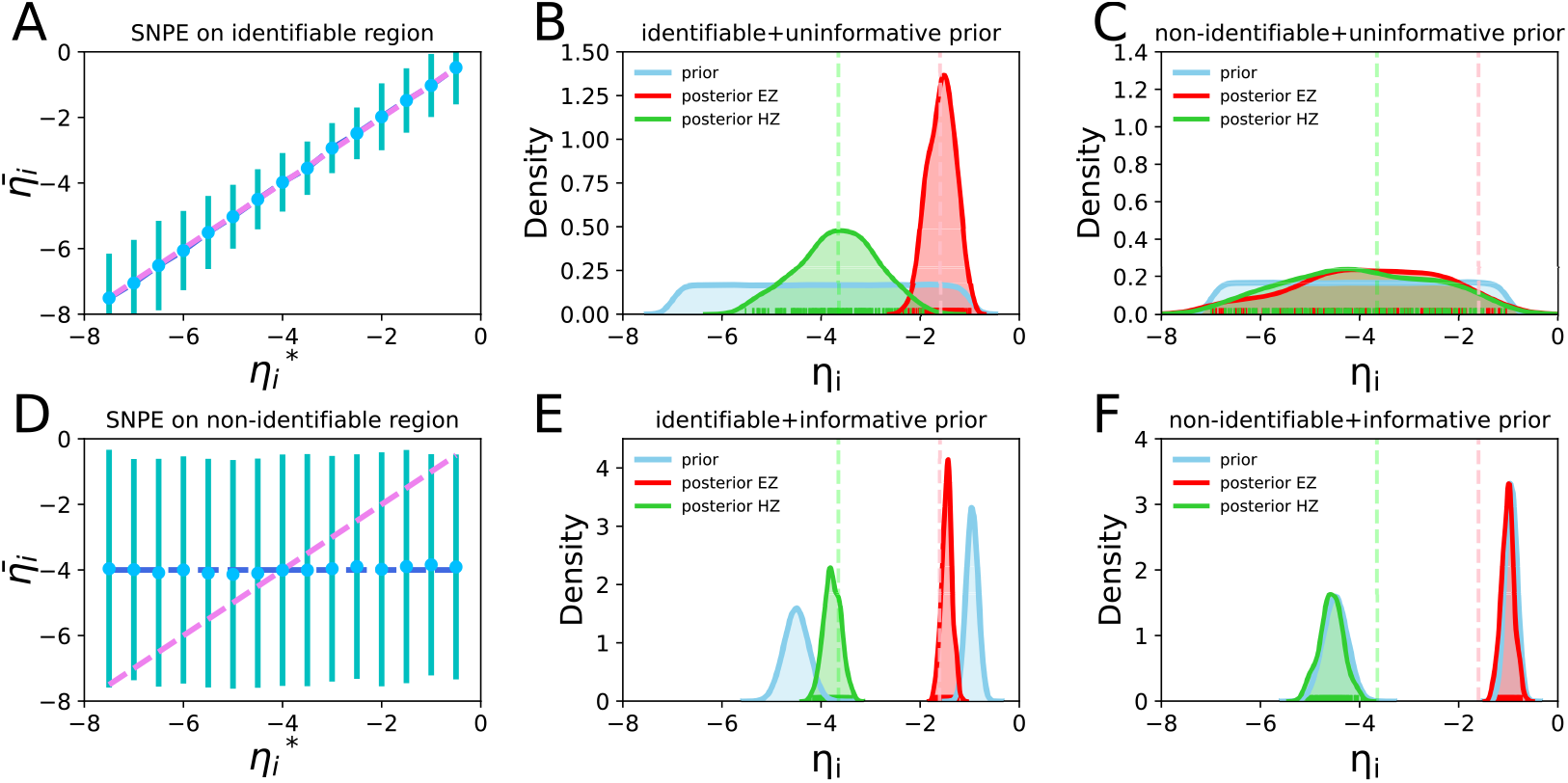
Bayesian identifiability analysis in the SBI-VEP. (**A**), (**D**) illustrate the uncertainty in the estimation using SNPE by varying the intrinsic dynamics in an identifiable and a non-identifiable region, respectively. Dashed magenta line represents a perfect fit. (**B**), (**E**) show the estimated posterior distributions by the use of uninformative and informative priors, respectively, for the identifiable regions. (**C**), (**F**) show the estimated posterior distributions by the use of uninformative and informative priors, respectively, for the non-identifiable regions. The green and red vertical dashed lines indicate the ground-truth for HZ and EZ, respectively.

Taking a Bayesian perspective, if the posterior distribution is affected by the changes in the prior setting, the parameter can be identifiable (prior predictive checking process). Fig. 6**B** and **E** demonstrate the estimated posterior distributions using uninformative and informative prior distributions, respectively, for identifiable regions (EZ and HZ close to electrodes, shown in red and green, respectively). It can be observed that the uninformative (diffuse) prior provides a reasonable amount of uncertainty for these regions (Fig. 6**B**), whereas the informative prior supplies less posterior variances than diffuse priors (Fig. 6**E**). This indicates that the resulting shrinkages in posteriors depend on the level of information (or variance) in the priors, thus they can be identifiable. Although the HZ was classified as practically non-identifiable by the likelihood-based approach, here, it drives finite confidence intervals in both upper and lower bounds due to the truncation in prior used for Bayesian setup. If the posterior distribution follows the prior with no shrinkage as shown in Fig. 6**C**, thus there is no relation between the posterior samples of such parameters, they are structurally non-identifiable (e.g., the distant regions from electrodes as their corresponding elements in the gain matrix are equal to zero). This problem can be addressed by the use of informative priors as shown in Fig. 6**F**. Although placing informative priors would mask the problem and bias the estimation, such information can be derived from complementary data such as lesions visible in the MRI or clinical evidence that suggests restrictions on the possible range of a particular parameter, or a relationship between parameters. Moreover, adding information through prior can dramatically improve the exploration of the search space in terms of computational cost and algorithmic diagnostics such as effective numbers of samples rendering the inference more efficient.

### 3.6. Algorithmic benchmark on the SBI-VEP

Our algorithmic benchmark consists of existing state-of-the-art approaches for neural network-based SBI namely SNLE, SNRE, and SNPE targeting the (weighted) log-likelihood, the likelihood ratios, or directly the posteriors, respectively. SNLE family utilizes deep neural networks to learn an approximation to the intractable likelihood followed by MCMC sampling to obtain the posterior. SNRE approaches use classifiers to learn likelihood ratios between two choices of parameters, and MCMC is used to obtain samples from the posterior. Rather, SNPE approaches use deep neural networks to directly target the posterior, bypassing the need for MCMC. Additionally, we use the current state-of-the-art implementations of Normalizing Flows (NFs) for training on simulations known as Mixture Density Network (MDN), Masked Autoregressive Flows (MAF), and Neural Spline Flows (NSFs). We ran these algorithms using SBI package (Tejero-Cantero et al., 2020), with the same model simulations in training data set, and observation for fitting. The metrics used here to rank the algorithmic performance are the accuracy in the estimation of EZ/PZ/HZ across whole-brain regions, the reliability of estimation (measured by posterior z-scores versus posterior shrinkages), and the computational cost for convergence.

We first ran different flow-based algorithms (MDN/MAF/NSFs) embedded in SNPE to infer excitability parameters across whole-brain regions at source-level measurement, in which all the parameters are identifiable (Fig. 7**A**). We observed that MDN failed to converge, whereas MAF and NSFs resulted in the same level of accuracy. Although NSFs provide more posterior shrinkages than MAF, it is 3 − 5 times slower for our model. The performance of MAF in the inference of spatial map of excitability parameters with different brain network sizes is also investigated for a toy model with *N*_*n*_ = 8, and *N*_*n*_ = 16 regions, only left hemisphere with *N*_*n*_ = 42, whole-brain using Desikan-Killiany parcellation with *N*_*n*_ = 84, VEP atlas with *N*_*n*_ = 162 and Schaefer parcellation with *N*_*n*_ = 400 regions. We observed that using seizure onset as the data feature with sufficient simulation budget, MAF scales to large brain network sizes (see Fig. 7**B**, and Fig. S8 for computational time involved in different steps of inference). Then, we embedded MAF in SNLE, SNRE, and SNPE families to infer the excitability parameters from sensor-level measurement, in which some of the brain regions are (structurally or practically) non-identifiable (Fig. 7**C**). Our results indicate that SNPE is more accurate than SNLE and SNRE to infer the spatial map of excitability parameters, also with proper amount of posterior shrinkage. SNLE showed the longest runtimes among the sequential algorithms and resulted in many false positive (type I) errors, whereas SNRE is the fastest but it is not able to provide proper posterior shrinkage (see Fig. S9). SNLE and SNRE rely on MCMC sampling to compute posteriors, and this sampling step can limit their performance. These results indicate that using MAF in SNPE to directly infer the posteriors is more well-suited for the VEP model (at both source and sensor levels) than alternative approaches (likelihoods or density ratios estimation) in terms of accuracy, the level of certainty in the estimation, and computational time for convergence.

**Fig. 7.**
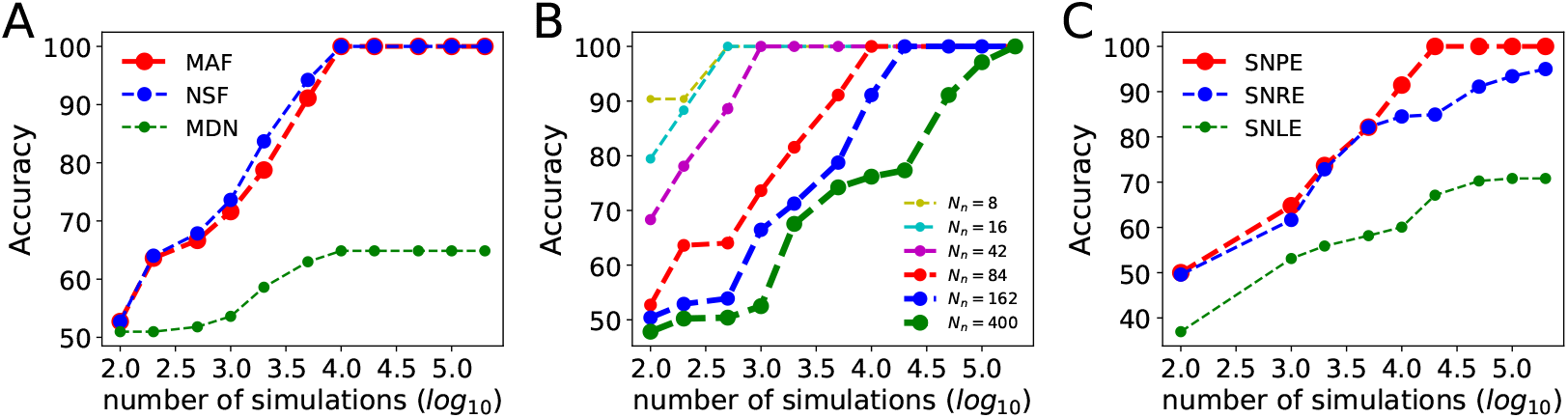
The state-of-the-art algorithms in the SBI-VEP. (**A**) Accuracy in inference from whole-brain regions at source-level using three flow-based algorithms MDN, MAF, and NSFs embedded in SNPE. (**B**) Accuracy in inference from whole-brain regions at source-level for different brain network sizes using MAF embedded in SNPE. (**C**) Accuracy in estimation from sparse SEEG recording at sensor-level using MAF embedded in three SBI approaches SNLE, SNRE, and SNPE.

### 3.7. The SBI-VEP against empirical SEEG recordings

Finally, the SBI-VEP is tested against a retrospective patient with drug-resistant right temporal lobe epilepsy (Engel score I, seizure-free). To this end, the constructed VEP model for this specific patient is fitted against the low-dimensional data feature of empirical SEEG recordings, using MAF in SNPE, and the inferred spatial map of epileptogenicity is compared with the clinical EZ hypothesis. The clinical classification is a binary classification labeling each region as part of EZ or not part of EZ, whereas the VEP model prediction-based classification consists of three classes as EZ/PZ/HZ. Here, clinical hypothesis about the EZ is incorporated in Bayesian inference as a weakly informative prior on the excitability parameter given by 𝒩 (−1.5, 1.0) for regions in EZ hypothesis, otherwise 𝒩 (−3.5, 1.0), where, 𝒩 represents a normal distribution. Figure 8**A** shows the fitting between the model prediction and observed data feature (total sensor power). A comparison of the fitted and observed SEEG envelope power is shown in Fig. S10. The inferred seizure initiation and propagation pattern for this patient is shown in Fig. 8**B**. Out of the six brain regions in the clinical EZ hypothesis, the model predicted three subcortical regions (r43: right-thalamus-proper, r47: right-hippocampus, r48: right-amygdala) in EZ where the seizure started, and three cortical regions (r54: ctx-rh-entorhinal, r55: ctx-rh-fusiform, r64: ctx-rh-parahippocampal) as part of PZ where only one of the regions (r55: ctx-rh-fusiform) recruited by the seizure. Fig. 8**C** and **D** illustrate the comparison of clinical EZ hypothesis with VEP model-predicted EZ/PZ. These results indicate that model predictions match well with the clinical hypothesis, while understanding the seizure propagation is not obvious unless accompanied by the inference on hidden source dynamics.

**Fig. 8.**
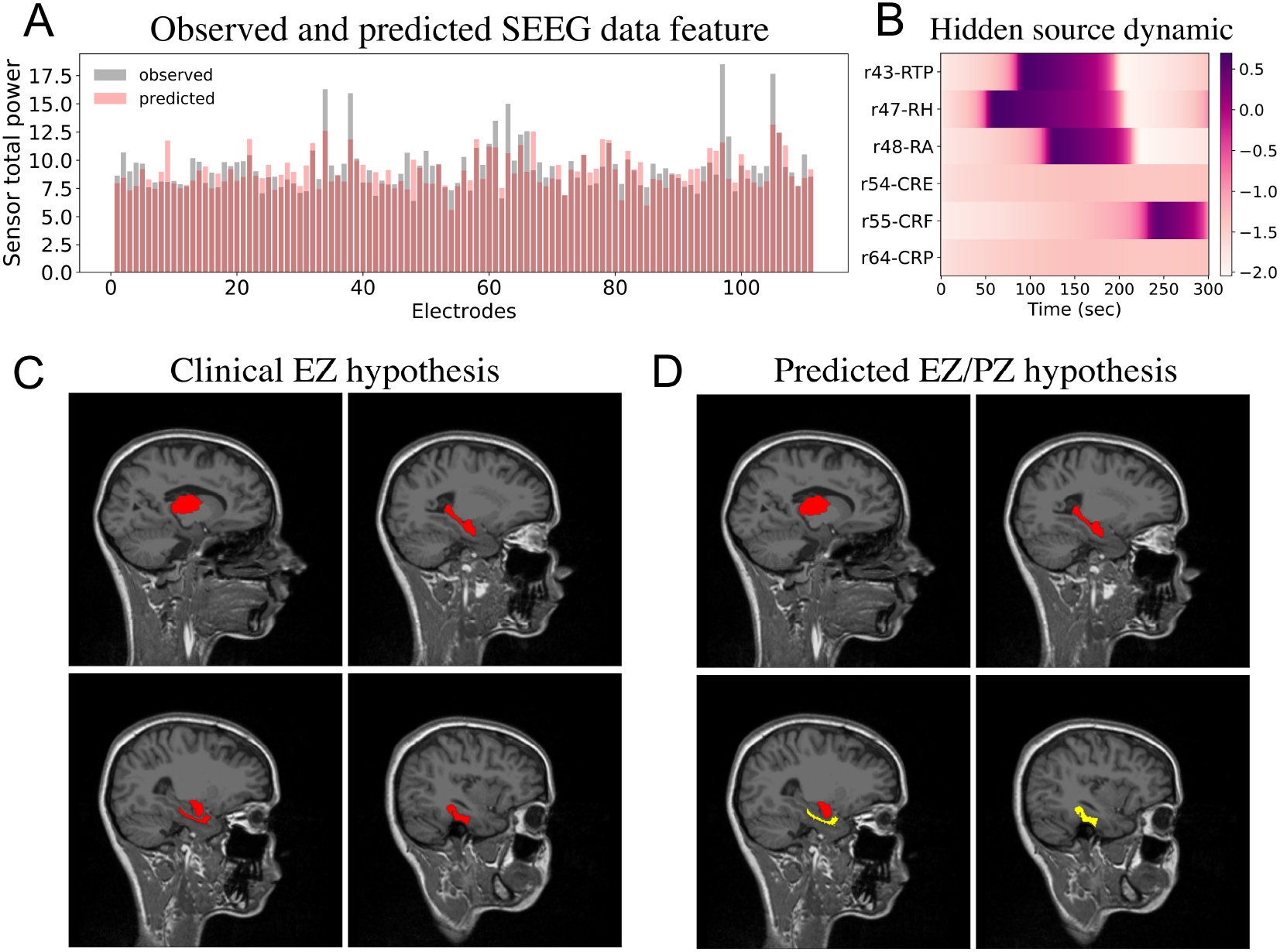
Inferred seizure initiation and propagation pattern from empirical SEEG recordings using SNPE. (**A**) Total sensor power as the data feature used to fit SEEG signals. Observations are shown in gray and model predictions are shown in red (RMSE=1.77). (**B**) Model-predicted seizure initiation and propagation pattern at (unobserved) source-level brain activity. (**C**) Clinical EZ hypothesis. (**D**) Model-predicted EZ/PZ hypothesis.

## 4. Discussion

In this study, based on a phenomenological model of seizure onset and propagation called Epileptor (Jirsa et al., 2014), a personalized large-scale brain model of epilepsy named as VEP (Jirsa et al., 2017) was constructed by placing Epileptor at parcelled brain regions, while each region is connected to other regions via the patient-specific connectivity matrix. Such a whole-brain modeling approach (Hashemi et al., 2020) combines the mean-field model of abnormal neuronal activity with the subject-specific brain’s anatomical information derived from non-invasive diffusion neuroimaging techniques (MRI, DTI). This allows us to simulate various spatio-temporal patterns of brain activity for a specific patient in a single computational framework. Then, a post-SEEG-implantation CT scan was used to find the exact locations of the SEEG electrodes and to construct the gain matrix which maps the simulated source activity to the corresponding SEEG signals at the sensors. However, identifying the spatio-temporal seizure propagation patterns has been a challenging problem due to the large dimensionality of the search space, non-linearity in system dynamics, uncertainty associated with unknown parameters and prior information, as well as the spatial sparsity of electrode implantation. More importantly, for model inversion at whole-brain scales such as VEP to identify the hypothetical areas, the calculation of likelihood function becomes intractable. In this study, we proposed an efficient Bayesian approach (SBI-VEP) to estimate the posterior distribution of spatial map of EZ/PZ in the brain, in which the methodology only requires forward simulations, rather than model-specific analytic calculation or exact evaluation of the likelihood function (Cranmer et al., 2020). We used state-of-the-art deep neural networks for density estimation in SBI families (Papamakarios and Murray, 2016; Lueckmann et al., 2017; Greenberg et al., 2019) to amortize the posterior distribution of EZ/PZ by training on low-dimensional data features extracted from a limited number of VEP model simulations (see Fig. 1). Using synthetic data, we demonstrated that the SBI-VEP is able to reliably infer the distribution of EZ/PZ from whole-brain activity at the source (see Fig. 2) and sensor (see Fig. 4) levels, while we needed fewer forward simulations having the seizure onset as the data features compared to time-invariant summary statistics and SEEG log power used for sparse SEEG data (e.g., 10 orders of magnitude for 84 brain regions). We also demonstrated that the SBI-VEP is able to accurately classify the stability of system fixed point at each brain region (EZ: unstable fixed point, PZ: close to bifurcation, and HZ: stable fixed point, shown in Fig. 3). Note that since we defined seizure propagation as a brain network effect, an accurate estimation of the global coupling parameter is critically necessary to avoid misclassification of PZ as EZ (by underestimation) or HZ (by overestimation). Importantly, SBI is able to deal with structurally and practically non-identifiable brain regions without impeding the inference on identifiable regions (see Fig. 5 and Fig. 6). This is a key advantage of SBI compared to gradient-based MCMC in which the convergence issues such as slow mixing and divergence appear in the presence of degeneracy (Betancourt et al., 2014; Betancourt, 2014b), making the entire sampling process inefficient. However, such algorithmic issues can be used as a powerful diagnostic tool to detect degeneracies (Betancourt, 2016a,b), whereas using SBI, it is required to additionally perform the identifiability analysis if there was no proper shrinkage in the posteriors. Nevertheless, how to find a reparameterization that reorganizes model configurations into spaces that are more consistent with the observed data for resolving degeneracy remains challenging.

It is known that approximate Bayesian computation (ABC) methods suffer from the curse of dimensionality and the choice of performance metric is critical (Cranmer et al., 2020). It has been shown that neural network-based approaches outperform ABC algorithms (Gonçalves et al., 2020), however, the performance ranking of these algorithms is task-dependent, and there is no uniformly best algorithm across different tasks (Lueckmann et al., 2021). Therefore, we performed a benchmark on expressive deep neural density estimators used in different approaches to SBI which do not require numerical evaluation of likelihoods, and thus can be used in the SBI-VEP framework. Among the current state-of-the-art algorithms for deep neural density estimator (MDN/MAF/NSFs) and SBI families that target the choice of likelihood, density ratios or directly posterior (SNLE, SNRE, SNPE, respectively), we found that using MAF in a single round SNPE is more accurate and efficient than alternatives in terms of overall accuracy in EZ/PZ estimation, the reliability of estimation, and the computational time involved in training and sampling (see Fig. 7). Notably, using MAF in SNPE scales to high-resolution brain parcellation (see Fig. 7), enables us to provide a higher predictive power in identifying the EZ/PZ, and thus better supporting surgery as a viable treatment option for drug-resistant epilepsy.

The SBI-VEP approach shares a key aspect with the well-established framework for inference in neuroimaging modalities called Dynamical Causal Modeling (DCM; Friston et al. (2003)): Bayesian inference over the parameters of a dynamical system model. However, there are key differences in practice. From modeling perspective, the SBI-VEP is a connectome-based approach that considers the structural connectivity (SC) as fixed parameters which are obtained from non-invasive imaging data of individuals. Whereas DCM is furnished with effective connectivity to explain the effects on observation induced by the actual changes in interactions among brain regions (i.e., a cause-effect relationship) (Frässle et al., 2017, 2018; Gilson et al., 2020; Prando et al., 2020), which can also better fit the model to empirical data. For instance, in the SBI-VEP, the seizure recruitment to a secondary subnetwork called PZ requires a strong connection with regions responsible for seizure initiation, the so-called EZ. Considering a data-driven approach such as tractography may limit the SBI-VEP capability to fit propagation to PZ that is weakly connected to EZ. In contrast, using effective connectivity, there are many more parameters to fit (e.g., *N* (*N* − 1)*/*2 symmetric connections for *N* regions), consequently, we will be able to capture the necessary connections for recruitment. However, inference on such a high-dimensional space can easily suffer from the non-identifiability issue, unless the changes in connectivity are constrained by the prior belief that there are transitions among a small number of brain connectivity states (Zarghami and Friston, 2020). The SBI-VEP considers non-linear brain dynamics at each brain region, whereas in DCM, the system dynamic is often linearized around its fixed point (on the state space equations of neuronal dynamics, and the observation equation for the measured signal) (Papadopoulou et al., 2015; Cooray et al., 2016). From the inference perspective, the SBI-VEP is equipped with state-of-the-art deep learning algorithms for statistical inference (i.e., estimating parameters of interest from observation given a model of their statistical relationship) and density estimation (i.e., estimating the joint probability density of a collection of random variables). The use of existing state-of-the-art algorithms in NFs such as MAF allows scaling the inference to whole-brain dynamics with high parcellation resolution (eg., *N*_*n*_ = 400 shown in Fig. 7). Notably, NFs used in the SBI-VEP approach systematically place a tighter bound on model evidence to deal with potential muti-modalities and degeneracies among parameters, whereas DCM is a variational-based approach and its mean-field variant ignores -by definition-the correlation between parameters.

Taking advantage of recent advances in probabilistic programming languages (PPLs) for automatic Bayesian inference such as Stan (a state-of-the-art platform for statistical modeling), the feasibility of inverting the coupled Epileptor model that best explains patient’s data from whole-brain source activity has been shown previously (Hashemi et al., 2020, 2021). In other studies, efficient and robust inversion of seizure propagation on whole-brain intracranial recordings were achieved by either simplifying the seizure dynamics using a threshold model (Sip et al., 2021) which considerably restricts the range of model dynamics, or by embedding Epileptor equations as priors on brain source dynamics and using maximum a posteriori (MAP) techniques, which does not capture the uncertainty in parameters (Vattikonda et al., 2021). These issues could possibly be addressed by sampling the whole posterior of Epileptor parameters using gradient-based and self-tuning MCMC sampling algorithms such as No U-Turn sampler (NUTS; Hoffman and Gelman (2014)). However, MCMC sampling of the posterior density from sparse observations such as SEEG recordings becomes computationally infeasible, in particular when parameters in a high-dimensional model such as VEP show strong non-linear correlations or if the posterior exhibits pathological geometries such as Neal’s funnel with varying curvature (Betancourt et al., 2014; Vattikonda et al., 2021). Recently, a hybrid of NFs (such as inverse autoregressive flows parameterized by neural networks) and HMC called neural-transport (NeuTra) HMC (Hoffman et al., 2019) has been proposed to correct this sort of unfavorable geometry, while it works in the amortized setting. The use of hybrid techniques like neural variational inference with gradient-based HMC sampler to achieve many of the benefits of HMC with much lower implementation and computational complexity, remains to be investigated and validated in future work. Another disadvantage of directly using the HMC in PPLs is the lack of amortization. The amortization strategy in SBI can be applied immediately to new data without repeated training. This allows us to rapidly evaluate different clinical hypotheses for a personalized model such as VEP, without computational overhead for further simulations.

Provided fast simulations, the methodology used in this study can be applied to other whole-brain models, since it requires neither model nor data features to be differentiable (Gonçalves et al., 2020). However, finding low-dimensional but sufficient informative data features that can deal with parameter degeneracies is the key challenge. The applications of SBI to estimate brain dynamics considering the existence of multiple fixed points, and the brain structure-function relationships with the parameter heterogeneity across regions affected by neurodegenerative diseases remain to be explored in future studies.

## Data Availability

The patient data sets cannot be made publicly available due to the data protection concerns regarding potentially identifying and sensitive patient information. Interested researchers may access the data sets by contacting Clinical Data Manager Aurelie Ponz at the Institut de Neurosciences des Systemes, Aix-Marseille Universite. The main source codes needed to reproduce the presented results are available on GitHub (https://github.com/ins-amu/BVEP).

## Abbreviations

VEP: virtual epileptic patient
TVB: the virtual brain
EZ: epileptogenic zone
PZ: propagation zone
HZ: healthy zone
SEEG: stereoelectroencephalography
MCMC: Markov chain Monte Carlo
HMC: Hamiltonian Monte Carlo
VI: variational inference
SBI: simulation-based-inference
ANNs: artificial neural networks
NFs: normalizing flows
MDN: mixture density network
NSFs: neural spline flows
MAF: masked autoregressive flow
SNPE: sequential neural posterior estimation
SNLE: sequential neural likelihood estimation
SNRE: sequential neural ratio estimation

## Information Sharing Statement

The patient data sets cannot be made publicly available due to the data protection concerns regarding potentially identifying and sensitive patient information. Interested researchers may access the data sets by contacting Clinical Data Manager Aurélie Ponz at the Institut de Neurosciences des Systèmes, Aix-Marseille Université. The main source codes needed to reproduce the presented results are available on GitHub (https://github.com/ins-amu/BVEP).

## Acknowledgements

This work was funded by the French National Research Agency (ANR; https://anr.fr) as part of the second “Investissements d’Avenir” program, ANR-17-RHUS-0004, EPINOV, to VJ and FB, by European Union’s Horizon 2020 Framework Programme for Research and Innovation under the Specific Grant Agreement No. 785907 and 945539, Human Brain Project SGA2 and SGA3 (https://ec.europa.eu/programmes/horizon2020) to VJ, by European Union’s Horizon 2020 Framework Programme for Research and Innovation under the Specific Grant Agreement and No. 826421, VirtualBrainCloud (https://ec.europa.eu/programmes/horizon2020) to VJ, PHRC-I 2013 EPISODIUM (grant number 2014-27), the Fondation pour la Recherche Médicale (DIC20161236442), and by SATT Sud-Est, 827-SA-16-UAM (https://www.sattse.com) to VJ and FB. The funders had no role in study design, data collection, and analysis, decision to publish, or preparation of the manuscript.

## Author contributions

M.H. and V.K.J. designed the study. F.B acquired the data. M.H. performed the study. M.H, A.N.V., J.J., V.S., M.M.W., F.B, and V.K.J. wrote the manuscript.

## Competing interests

The authors declare no competing interests.

## 5. Appendix

**Figure S1.**
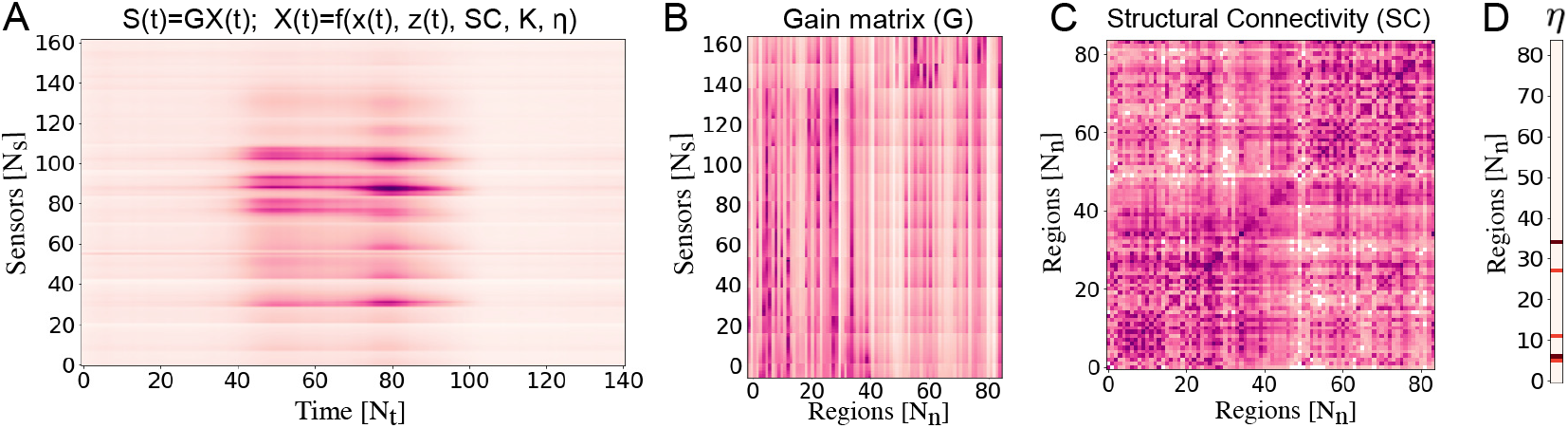
(**A**) The SEEG data modeled by *S*(*t*) = *GX*(*t*), where 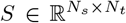 is sensor-level measurements, and 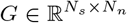 is gain matrix. The source brain activity 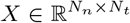 rendering as a non-linear function of hidden states *x*(*t*) coupled with slow variable *z*(*t*) given by Epileptor model, also the known structural connectivity 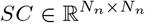, unknown global coupling parameter *K* ∈ ℝ^+^, and unknown brain regional parameter 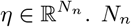: number of brain regions, *N*_*s*_: number of sensors, *N*_*t*_: number of data points per sensor. (**B**) The sparse gain (or lead-field) matrix of a randomly selected patient with partial epilepsy derived from the individual post-SEEG-implantation CT imaging data. Each element represents the inverse-squared distance between the region and the sensor. (**C**) Exemplary of SC, whose entries represent the connection strength between the brain regions, is derived from individual diffusion MRI tractography. Using Desikan-Killiany parcellation, the brain of the patient is parcellated into 84 different regions comprising 68 cortical regions and 16 subcortical structures. (**C**) The excitability (bifurcation) parameter *η*_*i*_ for each brain region. Colorbars: white to dark red correspond to low, respectively, high values.

**Figure S2.**
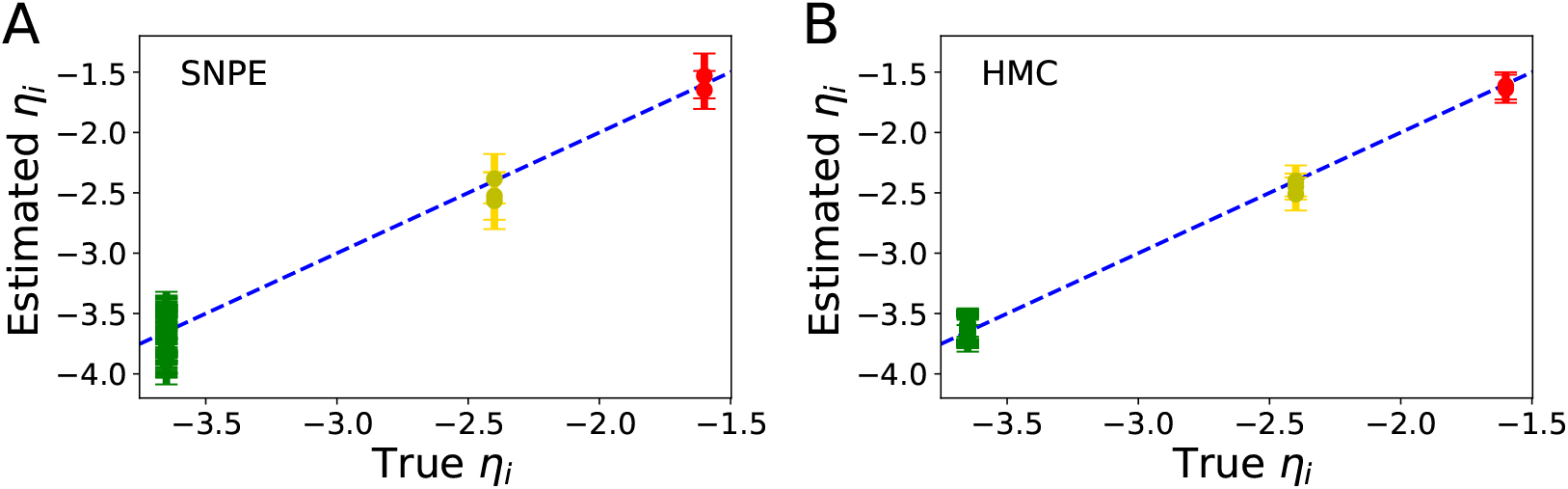
The estimated spatial map of epileptogenicity *η*_*i*_ for all brain regions at the sources of epileptic activity using (**A**) SNPE, and (**B**) HMC, versus the ground-truth used to generate the observation. Dashed blue line represents a perfect fit. The estimated *η*_*i*_ using SNPE are in a very close agreement with those obtained using HMC, in terms of accuracy with slightly more level of uncertainty, however, SNPE is significantly faster than HMC for the shown result. EZ: red, PZ: yellow, HZ: green.

**Figure S3.**
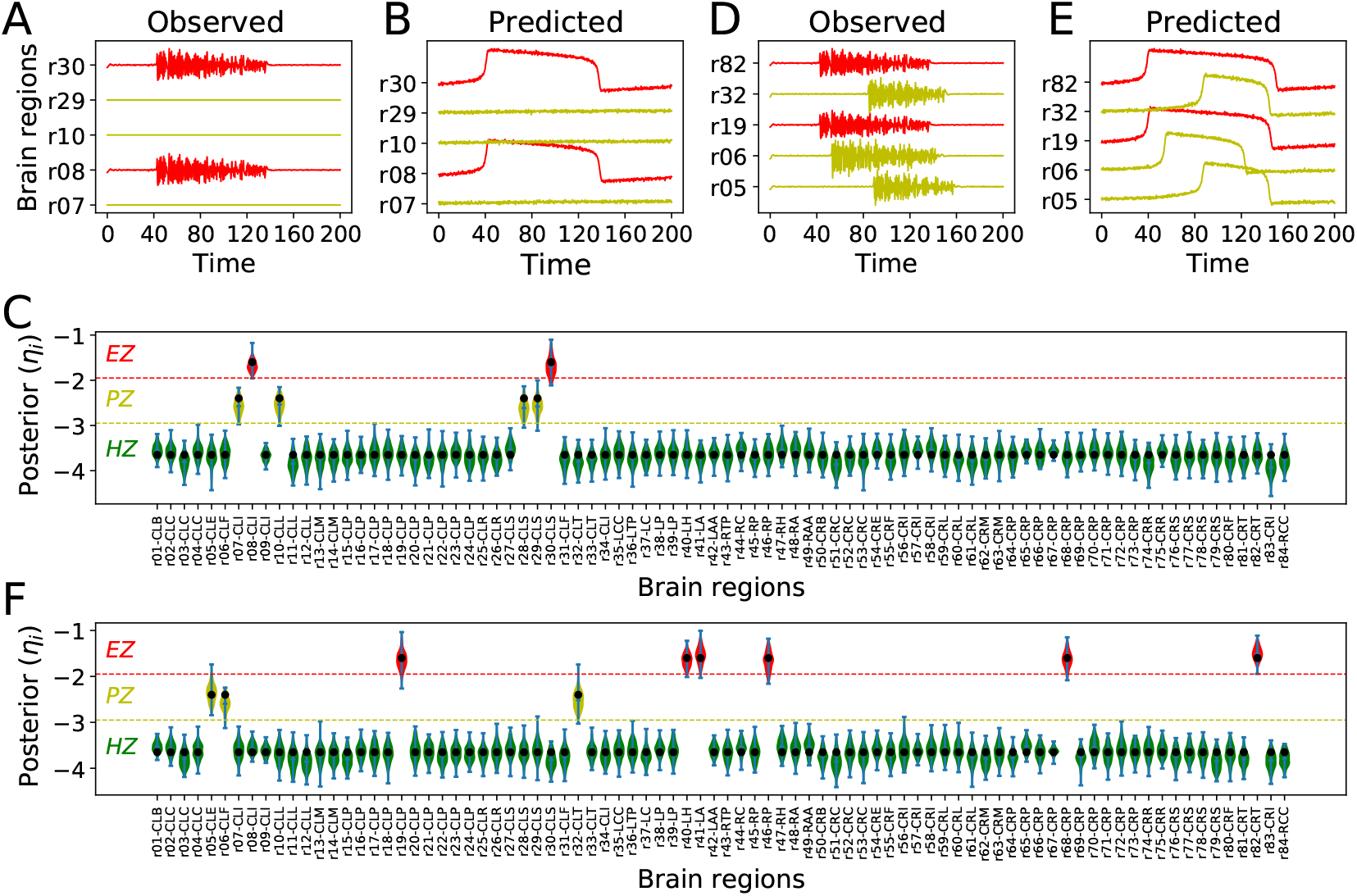
Amortized Bayesian inference of brain regional parameters at sources of epileptic activity. Due to the amortization of SNPE, an inference neural network is learned which can be applied to multiple data, enabling rapid inference on new EZ/PZ hypotheses without having to carry out new simulations for the fitting. (**A**), (**D**) The observed data generated by the full VEP model for two different spatial maps of epileptogenicity. (**B**), (**E**) The predicted data using SNPE trained on the reduced VEP model. The data features include statistical moments (*m* = 4) and the seizure onsets. (**C**), (**F**) Violin plots of the estimated posterior densities of excitability parameters. EZ: red, PZ: yellow, Ground-truth: filled black circles.

**Figure S4.**
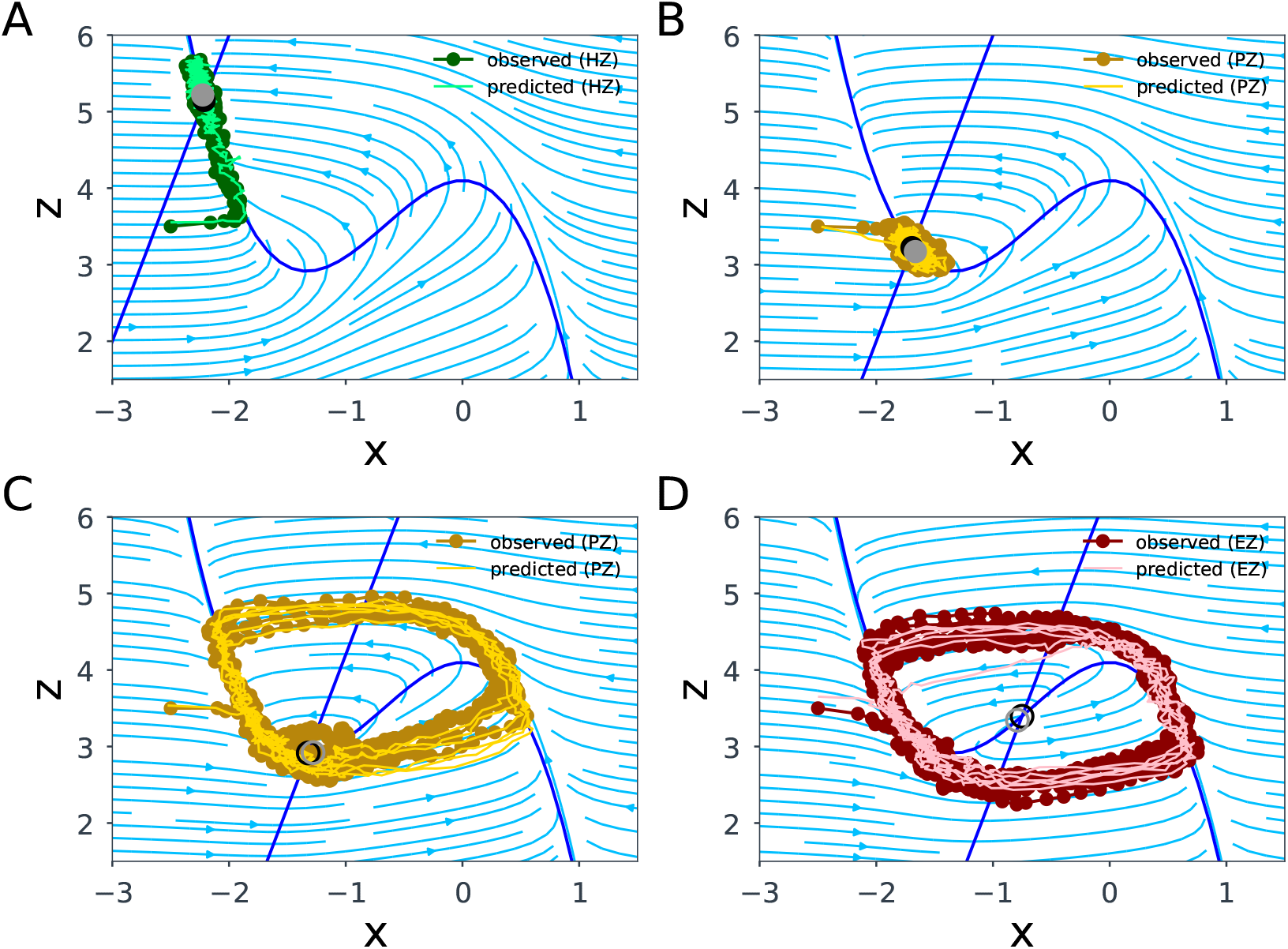
The observed and predicted 2D-VEP dynamics using SNPE trained on the system fixed point illustrated in the phase-plane for different brain regions. (**A**) HZ (not epileptogenic) exhibiting a single stable fixed point. (**B**) and (**C**) demonstrate the dynamics in PZ with coupling *K* = 0, and *K* = 1, respectively, as the candidate brain regions for seizure propagation. The equilibrium in PZ is close to the bifurcation value. (**D**) EZ exhibiting an unstable fixed point. The intersection of x- and z-nullclines (colored in blue) determines the fixed point of the system. Full circle and empty circle indicate the stable and unstable fixed points, respectively.

**Figure S5.**
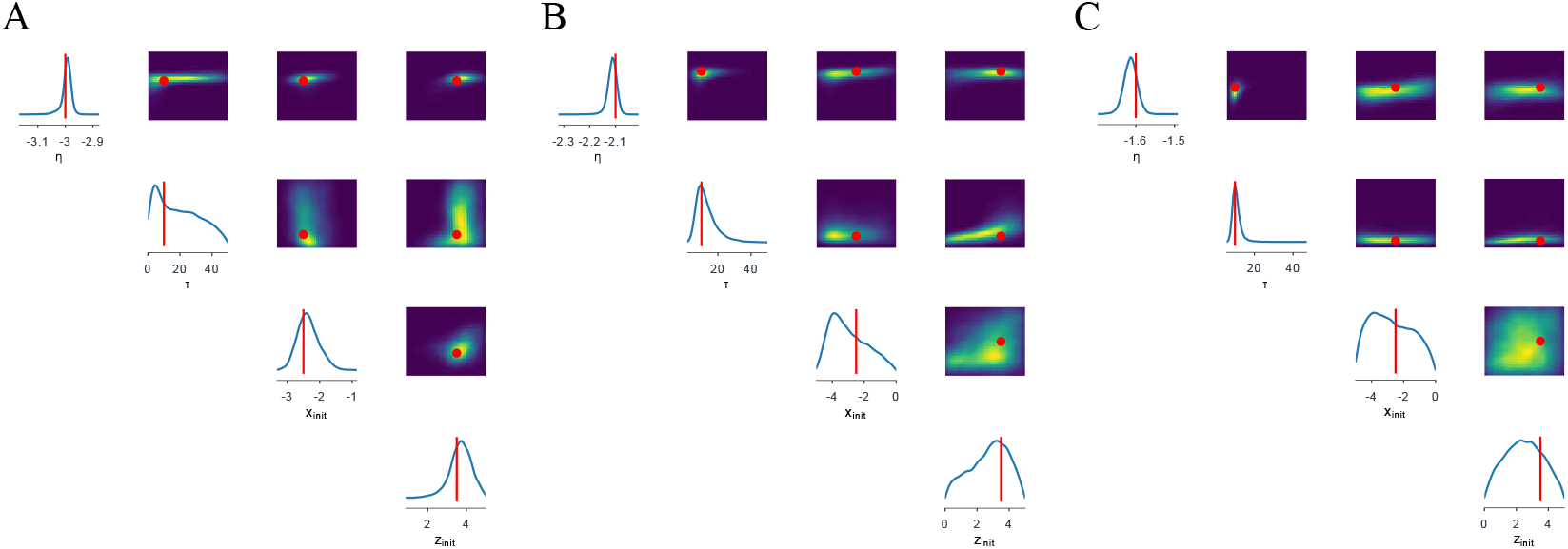
Estimated posterior and joint density distributions between Epileptor model parameters (excitability *η*, system time-scale *τ*, and initial condition of fast and slow variables *x*_*init*_ and *z*_*init*_) for different brain regions as (**A**) HZ, (**B**) PZ, and (**C**) EZ. SNPE with MAF accurately demonstrates the non-identifiability for the system time-scale in HZ, and the initial conditions in EZ and PZ, in agreement with the dynamical system properties. Ground truth parameters in red, high-probability in yellow, and low-probability in blue.

**Figure S6.**
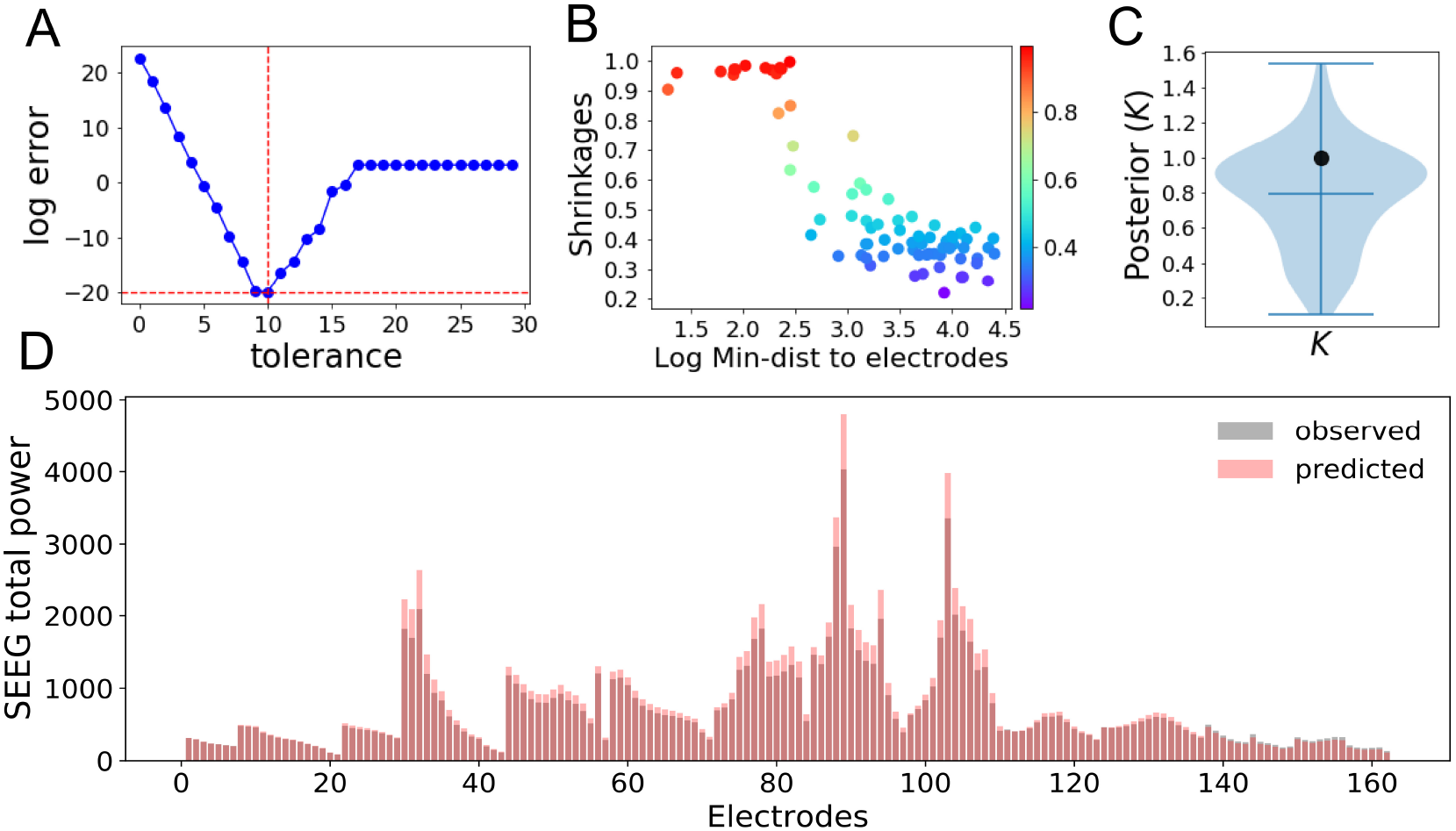
(**A**) Error between *Q* = *G*^⊤^*G* and *QQ*^−^*Q*, with G as the lead-field matrix, in the source localization using generalization (Moore-Penrose) method, indicating that the pseudo-inverse of matrix *Q* = *G*^⊤^*G* is very sensitive to the tolerance level, i.e., the cutoff for small singular values. Here, the source signal was estimated as *X* = (*G*^⊤^*G*)^−^*G*^⊤^*S*, where ⊤ denotes the matrix transposition, and (*G*^⊤^*G*)^−^ is g-inverse of *G*^⊤^*G* as *QQ*^−^*Q* = *Q*. (**B**) The posterior shrinkages using SNPE versus the minimum distance of brain regions from electrodes in log scale. (**C**) Posterior distribution of global coupling parameter *K* using SNPE. The prior was uniform truncated between *K* ∈ *𝒰* (0.0, 2.0). The filled black circles show the ground-truth used to generate the observed data. (**D**) Total sensor power as an informative data feature used to fit SEEG signals. Shown are the observed (in gray) and predicted (in red) SEEG envelope power using SNPE to estimate brain spatial parameters from 162 electrodes demonstrate a close agreement (RMSE=155.50).

**Figure S7.**
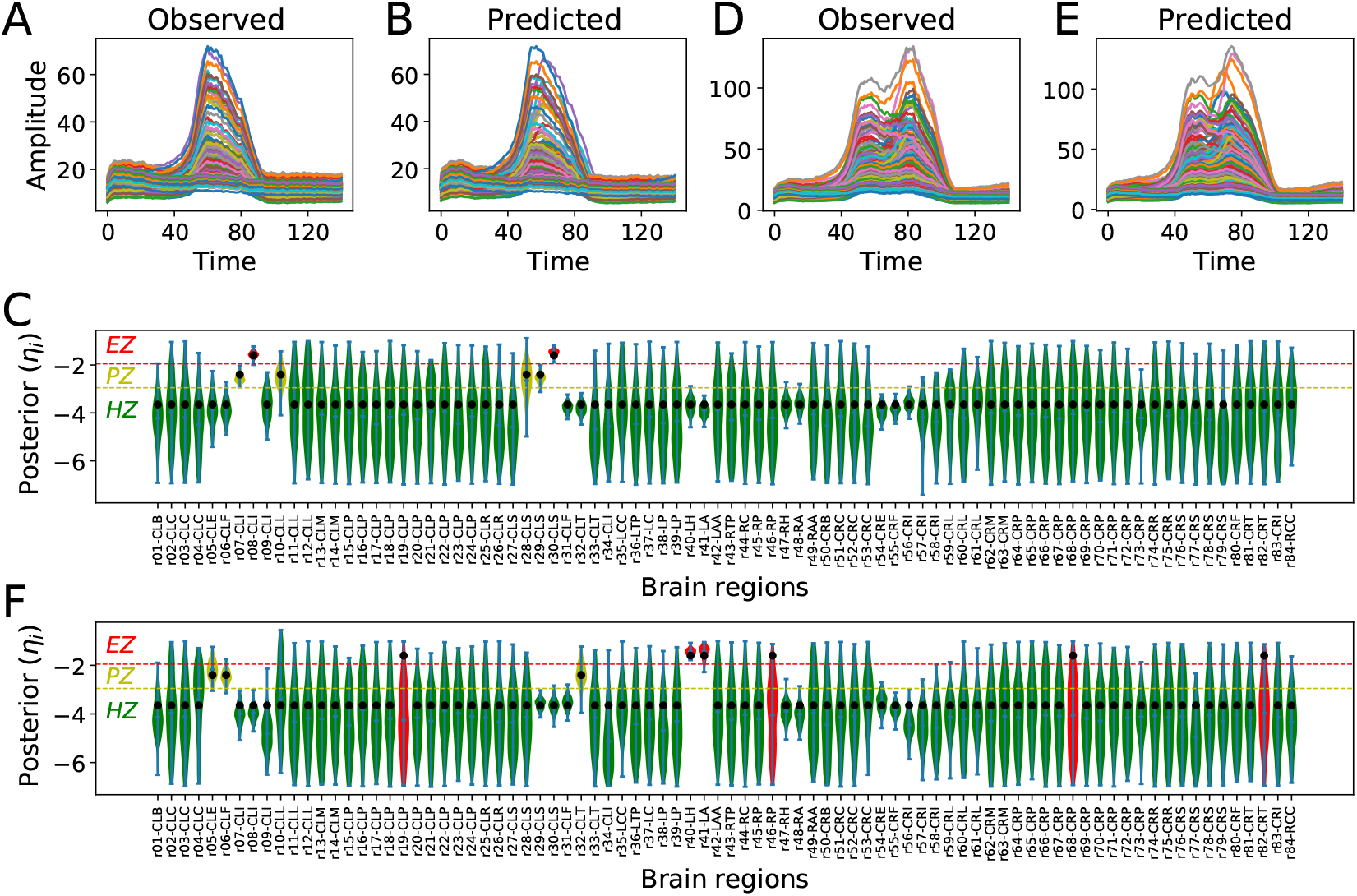
Amortized Bayesian inference of brain regional parameters from SEEG signals at sensor-level measurements. After the training, the amortization strategy in SNPE enables rapid inference on new EZ/PZ hypotheses without the need for the new simulation at fitting stage. (**A**), (**D**) The observed data generated by the reduced VEP model for two different spatial maps of epileptogenicity. (**B**), (**E**) The predicted SEEG data using SNPE. The data features include statistical moments (*m* = 4) and the seizure power (area under the signals). (**C**), (**F**) Violin plots of the estimated posterior densities of excitability parameters. EZ: red, PZ: yellow, Ground-truth: filled black circles.

**Figure S8.**
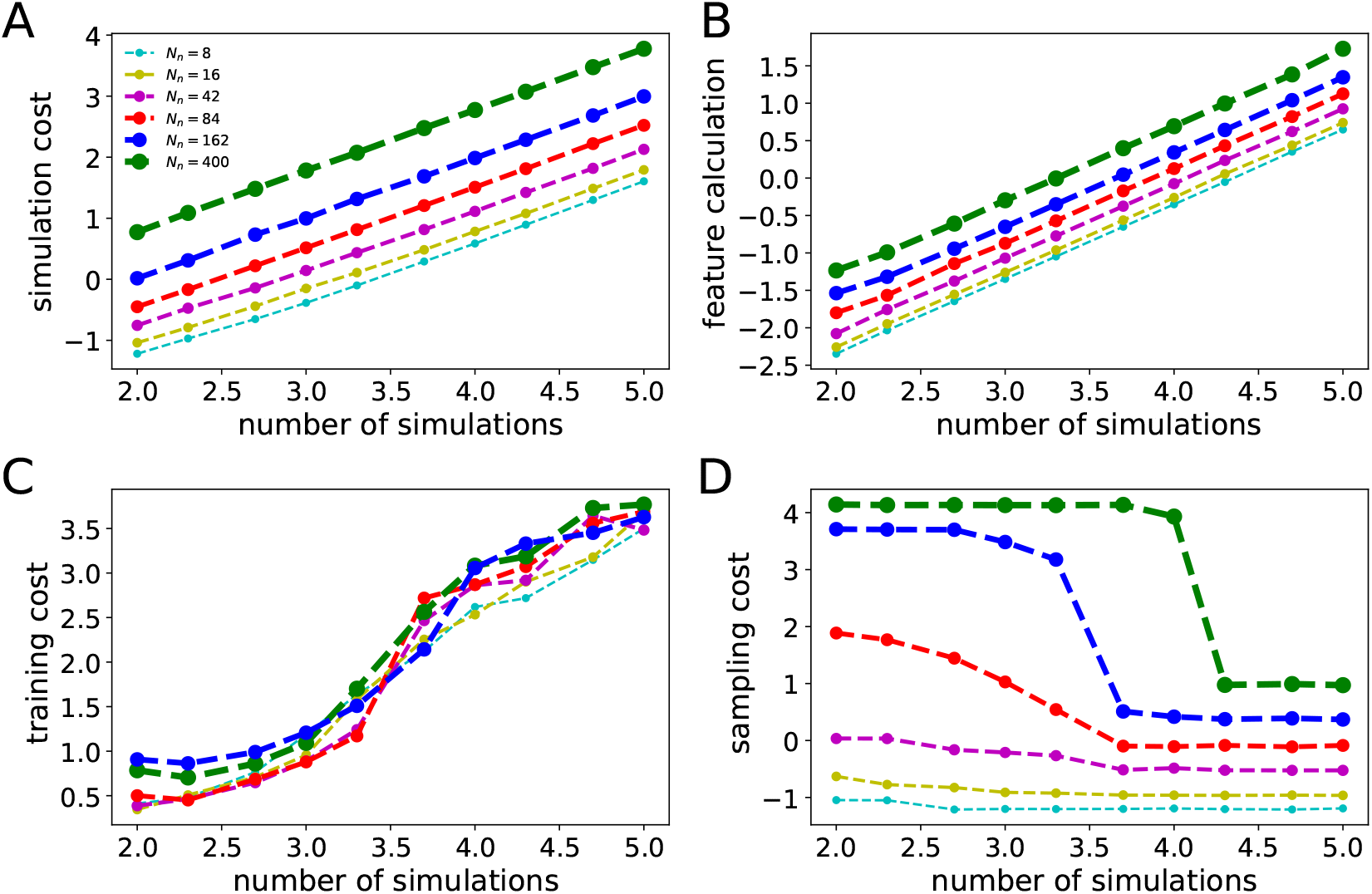
Computational time (in sec) involved in SBI-VEP with different brain network sizes, using MAF in SNPE, versus the simulation budget to obtain the posteriors. Here, the spatial map of excitability is inferred from the sources of epileptic activity with different brain parcellations (*N*_*n*_, the total number of brain regions, equivalently, the number of parameters to estimate), and only seizure onset is used as the data feature. A base-10 log scale is used on both the x- and y-axis. (**A**), (**B**) Model simulations using just-in-time (JIT) compilation, and calculation of data features in a single CPU, respectively, which both scale linearly with the number of simulations for training. (**C**) Training MAF in a single round SNPE scales exponentially with a number of simulations but is roughly independent from brain network size. (**D**) Sampling from the amortized posteriors depends on the number of brain regions (parameters), and it is slow with low number of simulations in training, but it becomes significantly faster for a sufficient number of simulations.

**Figure S9.**
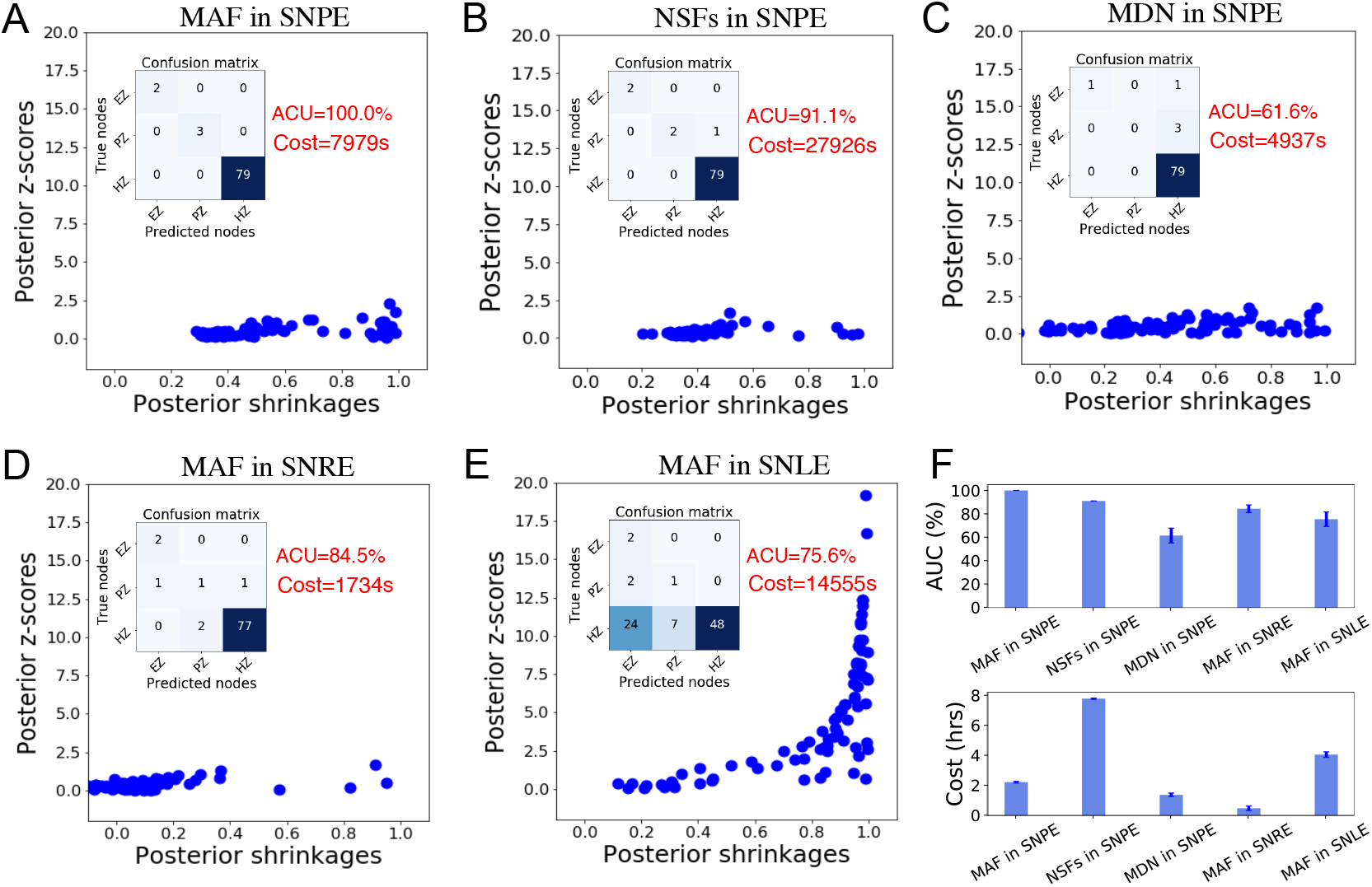
The state-of-the-art Normalizing Flows (MDN/MAF/NSFs) for training in SBI families (SNPE/SNLE/SNRE). (**A**) MAF in SNPE demonstrates the state-of-the-art performance in terms of accuracy in the estimation of spatial map of epileptogenicity (EZ/PZ/HZ) from sensor-level SEEG recordings, the reliability of estimation (measured by posterior z-scores versus posterior shrinkages), and computational cost for estimation. (**B**) NSFs is competitive with MAF in terms of accuracy and reliability, however, it is 3-5 times slower. (**C**) MDN is the fastest among Normalizing Flows, but it is not accurate and reliable according to posterior shrinkages. (**D**) SNRE is the fastest among SBI families, but it is not able to provide proper posterior shrinkages. (**E**) SNLE showed the longest runtimes among SBI families and resulted in many false positive (type I) errors. (**F**) Summarizes the accuracy and computational cost (training and sampling) averaged over four estimations.

**Figure S10.**
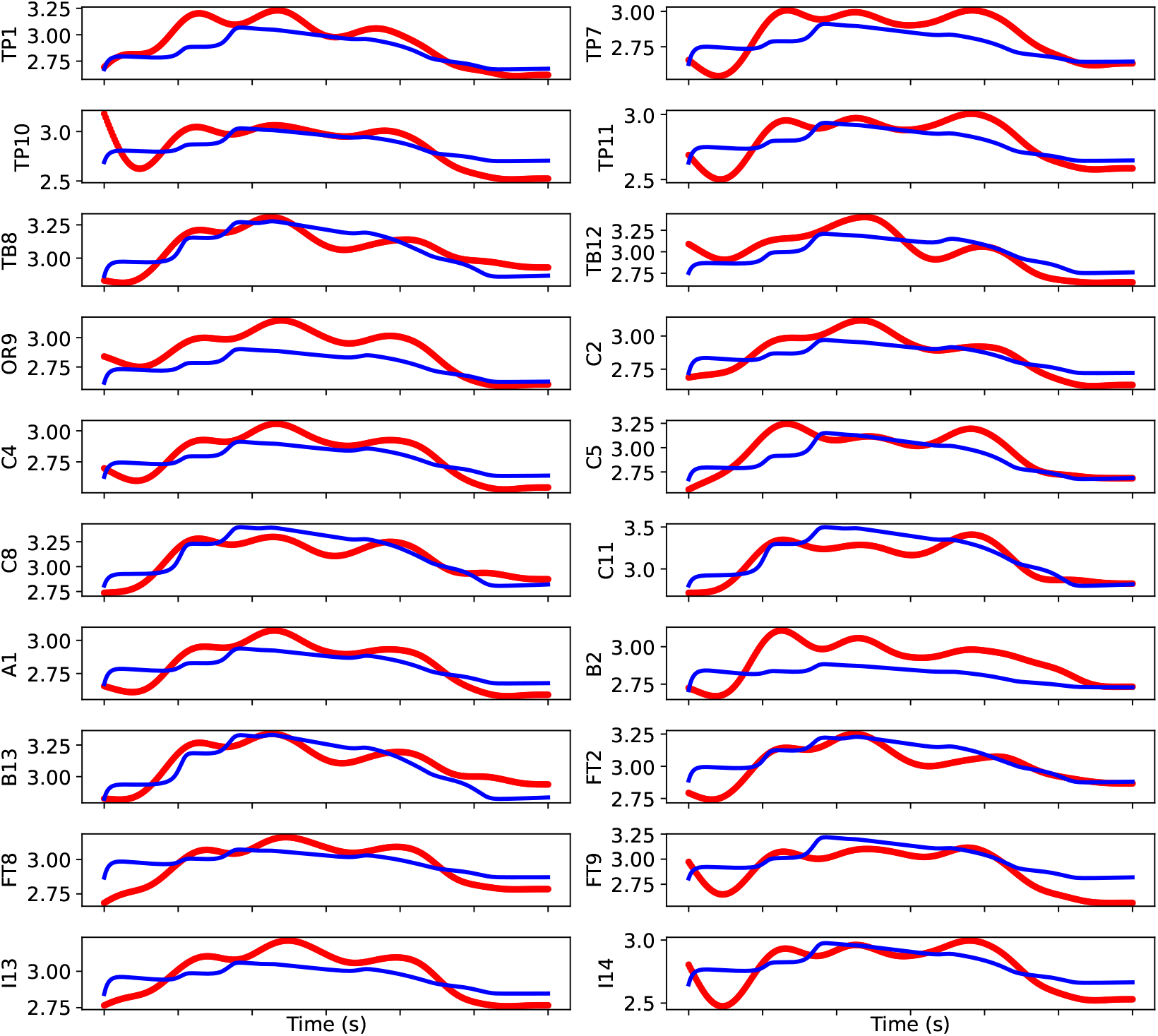
Comparison of model predicted (blue) with empirical (red) SEEG envelope power, using SNPE.

**Table S1.** Labels and indices of sub-divided brain regions.

| # | label | # | label |
| --- | --- | --- | --- |
| 01 | ctx-lh-bankssts | 43 | Right-Thalamus-Proper |
| 02 | ctx-lh-caudalanteriorcingulate | 44 | Right-Caudate |
| 03 | ctx-lh-caudalmiddlefrontal | 45 | Right-Putamen |
| 04 | ctx-lh-cuneus | 46 | Right-Pallidum |
| 05 | ctx-lh-entorhinal | 47 | Right-Hippocampus |
| 06 | ctx-lh-fusiform | 48 | Right-Amygdala |
| 07 | ctx-lh-inferiorparietal | 49 | Right-Accumbens-area |
| 08 | ctx-lh-inferiortemporal | 50 | ctx-rh-bankssts |
| 09 | ctx-lh-isthmuscingulate | 51 | ctx-rh-caudalanteriorcingulate |
| 10 | ctx-lh-lateraloccipital | 52 | ctx-rh-caudalmiddlefrontal |
| 11 | ctx-lh-lateralorbitofrontal | 53 | ctx-rh-cuneus |
| 12 | ctx-lh-lingual | 54 | ctx-rh-entorhinal |
| 13 | ctx-lh-medialorbitofrontal | 55 | ctx-rh-fusiform |
| 14 | ctx-lh-middletemporal | 56 | ctx-rh-inferiorparietal |
| 15 | ctx-lh-parahippocampal | 57 | ctx-rh-inferiortemporal |
| 16 | ctx-lh-paracentral | 58 | ctx-rh-isthmuscingulate |
| 17 | ctx-lh-parsopercularis | 59 | ctx-rh-lateraloccipital |
| 18 | ctx-lh-parsorbitalis | 60 | ctx-rh-lateralorbitofrontal |
| 19 | ctx-lh-parstriangularis | 61 | ctx-rh-lingual |
| 20 | ctx-lh-pericalcarine | 62 | ctx-rh-medialorbitofrontal |
| 21 | ctx-lh-postcentral | 63 | ctx-rh-middletemporal |
| 22 | ctx-lh-posteriorcingulate | 64 | ctx-rh-parahippocampal |
| 23 | ctx-lh-precentral | 65 | ctx-rh-paracentral |
| 24 | ctx-lh-precuneus | 66 | ctx-rh-parsopercularis |
| 25 | ctx-lh-rostralanteriorcingulate | 67 | ctx-rh-parsorbitalis |
| 26 | ctx-lh-rostralmiddlefrontal | 68 | ctx-rh-parstriangularis |
| 27 | ctx-lh-superiorfrontal | 69 | ctx-rh-pericalcarine |
| 28 | ctx-lh-superiorparietal | 70 | ctx-rh-postcentral |
| 29 | ctx-lh-superiortemporal | 71 | ctx-rh-posteriorcingulate |
| 30 | ctx-lh-supramarginal | 72 | ctx-rh-precentral |
| 31 | ctx-lh-frontalpole | 73 | ctx-rh-precuneus |
| 32 | ctx-lh-temporalpole | 74 | ctx-rh-rostralanteriorcingulate |
| 33 | ctx-lh-transversetemporal | 75 | ctx-rh-rostralmiddlefrontal |
| 34 | ctx-lh-insula | 76 | ctx-rh-superiorfrontal |
| 35 | Left-Cerebellum-Cortex | 77 | ctx-rh-superiorparietal |
| 36 | Left-Thalamus-Proper | 78 | ctx-rh-superiortemporal |
| 37 | Left-Caudate | 79 | ctx-rh-supramarginal |
| 38 | Left-Putamen | 80 | ctx-rh-frontalpole |
| 39 | Left-Pallidum | 81 | ctx-rh-temporalpole |
| 40 | Left-Hippocampus | 82 | ctx-rh-transversetemporal |
| 41 | Left-Amygdala | 83 | ctx-rh-insula |
| 42 | Left-Accumbens-area | 84 | Right-Cerebellum-Cortex |

## Notes

### Competing Interest Statement

The authors have declared no competing interest.

### Author Declarations

Informed written consent was obtained for all patients in compliance with the ethical requirements of the Declaration of Helsinki and the study protocol was approved by the local Ethics Committee (Comite de Protection des Personnes sud Mediterranee 1).

